# Is more better? An analysis of toxicity and response outcomes from dose-finding clinical trials in cancer

**DOI:** 10.1101/2020.08.18.20177212

**Authors:** Kristian Brock, Victoria Homer, Gurjinder Soul, Claire Potter, Cody Chiuzan, Shing Lee

## Abstract

The overwhelming majority of dose-escalation clinical trials use methods that seek a maximum tolerable dose, including rule-based methods like the 3+3, and model-based methods like CRM and EWOC. These methods assume that the incidences of efficacy and toxicity always increase as dose is increased. This assumption is widely accepted with cytotoxic therapies. In recent decades, however, the search for novel cancer treatments has broadened, increasingly focusing on inhibitors and antibodies. The rationale that higher doses are always associated with superior efficacy is less clear for these types of therapies. We extracted dose-level efficacy and toxicity outcomes from 115 manuscripts reporting dose-finding clinical trials in cancer between 2008 and 2014. We analysed the outcomes from each manuscript using flexible non-linear regression models to investigate the evidence supporting the monotonic efficacy and toxicity assumptions. We found that the monotonic toxicity assumption was well-supported across most treatment classes and disease areas. In contrast, we found very little evidence supporting the monotonic efficacy assumption. Our conclusion is that dose-escalation trials routinely use methods whose assumptions are violated by the outcomes observed. As a consequence, dose-finding trials risk recommending unjustifiably high doses that are harmful to patients. We recommend that trialists consider experimental designs that allow toxicity and efficacy outcomes to jointly determine the doses given to patients and recommended for further study.

## 1 Introduction

A goal of dose-finding clinical trials is to evaluate outcomes under a set of investigational doses. A common general approach starts by giving a relatively low dose to a small cohort of patients. The outcomes of this cohort affect the dose given to the next. For instance, if no unacceptable toxic reactions are seen in this initial cohort, the next cohort is likely to be given a higher dose. However, if outcomes are adverse, the next cohort might be given the same dose or a lower dose, or the trial might be halted altogether. This sequential and adaptive approach continues until the experimental design identifies a suitable dose. This pattern of starting low and seeking to escalate dose justifies the descriptor *dose-escalation trials*.

The most common approach [1, 2] in dose-escalation trials is the family of rule-based A+B designs, the most famous example of which is the perennial 3+3 design [3]. It evaluates doses in cohorts of three patients, using a set of rules to escalate dose so long as an unacceptably high incidence of dose-limiting toxicity (DLT) is not seen.

The main alternative class of dose-escalation methodology comprises the so-called model-based designs. These use statistical models to estimate the dose-event curve. Whilst model-based methods are used far less frequently than rule-based methods, two designs that have seen relatively wide use in recent years [2] are the continual reassessment method (CRM) [4] and the escalation with overdose control (EWOC) method [5].

Biostatisticians have encouraged trialists to shift from rule-based to model-based methods, largely because they offer better performance [6–8]. The two approaches, however, share some fundamental assumptions. Firstly, they each assume that the probability of DLT increases as dose is increased. This reflects the toxicologists’ adage *the dose makes the poison*, a rule that is generally accepted without challenge. The 3+3, CRM and EWOC designs all select doses based only on binary DLT outcomes in pursuit of the maximum tolerable dose (MTD), the highest dose with toxicity probability less than some critical prespecified value. So long as two doses are deemed tolerable, the higher dose is favoured. Efficacy outcomes like response or survival are not formally used in dose-selection decisions. There are variants of the CRM design [9, 10] and other statistical approaches [11–13] for dose-finding by co-primary toxicity and efficacy outcomes. However, these have experienced comparatively little use [2, 14].

In dose-finding trials, investigators seek to identify tolerable and efficacious doses. The reliance solely on a toxicity outcome in the majority of dose-finding trials dictates that an assumption is made about the efficacy outcome.

*The assumption is that higher doses are always more efficacious*.

We refer to this as the monotonic efficacy assumption. In MTD-seeking trials, investigators will escalate dose without formal reference to an efficacy outcome. When the monotonic efficacy assumption holds, the MTD maximises the expectation of efficacy for a specified risk of toxicity. However, a plausible way in which the monotonic efficacy assumption can be violated is when the probability of efficacy plateaus at some critical dose. Escalation beyond this point exposes patients to a greater risk of toxicity for no accompanying increase in the probability of efficacy. As such, the monotonic efficacy assumption is rather more controversial than the monotonic toxicity assumption.

Monotonic efficacy has a plausible rationale in the treatments that have formed the backbone of anti-cancer therapies for decades. Cytotoxic treatments like chemotherapy damage tumours and healthy tissue alike. The presence of toxicity is a sign that anti-tumour activity is probably happening as well. In this setting where toxicity and efficacy are broadly accepted to be coincident, the use of dose escalation designs became commonplace. In recent years, however, numerous targeted therapies, immunotherapies and cell therapies have been approved for use against cancer. In a recent systematic review of dose-finding methodologies used between 2008 and 2014, Chiuzan *et al*. [2] found that over half of the trials investigated a targeted therapy or immunotherapy and the overwhelming majority used an MTD-seeking method. With these novel treatment classes, the rationale for assuming that efficacy always increases in dose is less clear.

There are notable instances in the literature where a monotonic dose-efficacy relationship has not been observed. For instance, a major recent success in the development of novel anti-cancer drugs has been the PD-1 blockade antibody, pembrolizumab. A phase I trial investigated pembrolizumab doses of 1 mg/kg, 3 mg/kg and 10 mg/kg every 2 weeks, and 2 mg/kg and 10 mg/kg every 3 weeks in 30 patients with various malignancies [15]. Large expansion cohorts in non-small-cell lung cancer (NSCLC) further evaluated 495 patients at doses 2 mg/kg or 10 mg/kg every 3 weeks or 10 mg/kg every 2 weeks [16]. Subsequently, a phase III trial that contributed to a licensing application in NSCLC randomised 345 patients to pembrolizumab 2 mg/kg, 346 to pembrolizumab 10 mg/kg, and compared each of these experimental arms to a control arm comprising 343 patients randomised to docetaxel [17]. Despite the wide range of doses investigated in large sample sizes, the phase III trial observed very similar overall survival and RECIST response outcomes in the two pembrolizumab doses, with each producing materially better outcomes than the control arm. The drug was subsequently licensed at 200 mg (i.e. not adjusted for patient weight) every 3 weeks, reflecting the absence of extra efficacy at higher doses. Assuming an average adult weight of 70–80kg, the licensed dose is situated at the lower end of the doses investigated throughout these three clinical trials.

We naturally wonder about the suitability of the monotonic efficacy assumption in a wider sense. In this manuscript, we investigate two related questions. What evidence is there that the probabilities of a) toxicity, and b) efficacy increase in dose in modern cancer therapies?

## 2 Methods

We sought to identify a broad sample of manuscripts reporting recent dose-finding clinical trials in oncology.

### 2.1 Identifying manuscripts

Chiuzan *et al*. [2] conducted a systematic review of the methods used in cancer dose-finding trials. Their findings mirrored those of Rogatko *et al*. [1] from the previous decade that over 90% of dose-finding trials use a rule-based design like 3+3. The authors found 1,712 manuscripts published between 2008 and the first half of 2014. The authors published in their paper a large table summarising the trials that used model-based methods, like CRM or EWOC, of which there were 92 examples. Whilst extracting data from 1,712 papers would be infeasible, extracting data from 92 was possible. However, the subset of trials that use model-based methods may not be representative of the entire sample. For this reason, we supplemented the list of 92 model-based papers with 30 randomly-selected papers that used rule-based methods, stratified by year of publication. Combined, this produced a sample of 122 manuscripts.

### 2.2 Extracting data

Each of the 122 manuscripts[18–139] presented the results of at least one dose-finding experiment in humans. Some papers reported the results of more than one experiment. From each experiment, we extracted descriptive data pertaining to the patient population, the dose-varying treatment, and any concomitant treatments. Concerning outcomes, we extracted the dose-levels administered, the number of patients evaluated at each, and the number of DLT and objective response events recorded at each. These outcomes are explained and justified in the following sections.

#### 2.2.1 Dose-level outcomes vs pooled outcomes

We only recorded outcomes broken down by dose-level because these would allow us to address the research question pertaining to the evidence for monotonically increasing toxicity and efficacy probabilities. We did not collect outcomes that were reported by pooling all dose-levels because this would not address our research question.

#### 2.2.2 Toxicity outcomes

Dose-limiting toxicity (DLT) is the de-facto standard safety outcome in dose-finding trials. Manifestation of DLT involves the occurrence of pre-specified adverse events (AEs) that are serious enough that they would motivate the clinicians to rule out higher doses in the affected patient or consider the temporary suspension or complete cessation of current therapy. There is no standard definition of DLT across trials but the outcome would be defined in each trial protocol and remain consistent across the doses investigated within each trial. The definition of DLT in a given trial may reflect the clinical characteristics of the disease and the anticipated adverse events from the entire treatment ensemble (i.e. arising from the experimental therapy and concomitant therapies).

Data on DLT outcomes were sought in every manuscript. We analyse outcomes for DLT because it was the most widely-reported toxicity outcome measure.

#### 2.2.3 Efficacy outcomes

The question motivating this research concerns drug efficacy and how this changes as dose is increased. *Efficacy* is only loosely defined in cancer. There is no single outcome that is unambiguously accepted as the variable best reflecting efficacy. Applications for drug licensing are generally supported by phase III trials that use survival outcomes like overall survival (OS) and progression-free survival (PFS). In contrast, early phase trials, when they evaluate efficacy, tend to use surrogate outcomes that can be evaluated over the short-term like disease response.

Assessing disease response generally involves comparing the extent of disease (e.g. tumour size or number of leukaemic cells) at baseline and after treatment administration to characterise the patient’s response to treatment using one of several categories. RECIST [140] is the most common response outcome categorisation used in solid tumour trials. RECIST categorises each disease assessment as one of: complete response (CR); partial response (PR); stable disease (SD); or progressive disease (PD).

Researchers have defined analogues to RECIST in other cancers, including blood cancers where diseased cells reside in the blood rather than a discrete measurable mass. An example of this is the Cheson criteria in acute myeloid leukaemia (AML) [141] and iwCLL criteria in chronic myeloid leukaemia [142]. These contain response categories that are similar to those in RECIST, with slight modifications to reflect the phenomena specific to the disease.

Under RECIST, an objective response (OR) is said to occur when a patient experiences CR or PR. Under the RECIST analogues, further response categories are included in OR. For instance, in AML, a patient with complete remission with incomplete blood count recovery would be considered to have experienced OR.

Data on OR outcomes were sought in every manuscript. We analyse outcomes for OR because it was the most widely-reported efficacy outcome measure.

#### 2.2.4 Orderability of doses

Analysing how the probabilities of events change as dose increases requires that we are working with increasing doses. The general 3+3, CRM and EWOC methods require that the doses under investigation are *fully orderable*. That is, we need to be able to unambiguously say that *d_i_ < d_j_* or *d_i_ > d_j_* for each pair of doses in the set of doses under investigation.

When we encountered dose-levels that were not fully orderable, for the purposes of conducting statistical analysis we broke the doses up to form fully orderable subsets that we called *analysis series*.

There are many possible subsets of a set so the way we formed the analysis series was unavoidably subjective. To promote objectivity, we followed some simple rules. We sought to maximise the size of the largest fully orderable series. Furthermore, we avoided allocating a dose to several series unless repetition was the only way to avoid having an orphan dose (i.e. a series of size 1).

Consider, for instance, the three dose scenario: dose 1 = 10mg of drug A + 20mg of drug B; dose 2 = 20mg A + 10mg B; dose 3 = 20mg A + 20mg B. This set of doses is not totally orderable because it is impossible to say whether dose 1 is higher or lower than dose 2. However, each of these doses is categorically less than dose 3. Thus, in this hypothetical scenario, we would have analysed outcomes of the two totally orderable subsets (dose 1, dose 3) and (dose 2, dose 3). In doing so, the outcomes at dose 3 would have featured in the analyses twice. The alternative would have been to arbitrarily throw away the outcomes at dose 1 or dose 2, an option we rejected because it is wasteful.

In summary, the data have been recorded in a way amenable to answering the research questions.

#### 2.2.5 Database creation

Data were extracted from papers and recorded on prior-written standardised forms. The data were then recorded on sheets in an Excel file that was deposited in the University of Birmingham’s data repository. [143]

Data were extracted by VH, GS, KB and CP. Data for 52 manuscripts were extracted by two different authors and differences were resolved by discussion. Data for 70 manuscripts were extracted by one author.

### 2.3 Analysis model

The DLT and OR outcomes we extracted were binary in nature. Outcomes were analysed within study using Emax models [144].

Emax is a flexible non-linear approach for fitting sigmoidal (i.e. S-shaped) dose-response curves to continuous or binary responses. In our analysis, the binary response variable was the patient-level presence of DLT or OR. The explanatory variable was the dose-level administered. Emax can capture relationships where the mean response increases in dose, decreases in dose, or is independent of dose. Separate models were fit to the collection of patient outcomes in each study. Outcomes from different studies were not pooled because of the disparate patient populations and definitions of DLT and OR. Each of these factors remained consistent within each analysis series. The fitted values from the Emax models represent the event probabilities at each dose, ranging from 0 to 1.

Binary outcomes might generally suggest analysis via logit linear regression models. We did not use logit models because they assume that the event probability invariably tends to 1 with large enough predictor values. This is inappropriate in our analysis that seeks to investigate how event probabilities vary with dose, including the possibility that event probabilities plateau at values less than 1. Whilst it may be acceptable to assume that a high enough dose will guarantee a toxic outcome, empirically it is inappropriate to assume that a high enough dose guarantees an efficacious response. The relative strength of the Emax model is that it allows the event probability to plateau at a value less than 1. It contains as a special case the scenario reflected by logit models where the event probability tends to 1 as dose is increased.

We used both maximum likelihood and Bayesian approaches to fit Emax models. Maximum likelihood models were fit because they do not require the specification of priors. However, the analysis series in this research were very small, with some data-sets including fewer than 10 patients. In several instances, maximum likelihood models failed to converge. In these circumstances, Bayesian models can be extremely valuable because the specification of very small amounts of information in prior distributions promotes model convergence. Bayesian models succeeded in estimating dose-event curves in all instances. Our chosen prior distributions are introduced briefly below and expanded in detail in the supplementary appendix.

The height of each dose-event curve was calculated as the model-fitted event probability at the highest dose under investigation minus that of the lowest dose. This concept is illustrated graphically in Figure 5 in the supplementary appendix.

#### 2.3.1 Prior distributions

For the Bayesian analyses, we were required to specify prior distributions on the four parameters in the Emax model. We selected uniform priors on the parameters that reflect the minimum and maximum event probabilities, constrained to take values only in the region from 0 to 1. This meant that all event probabilities at the lowest and highest doses were equally likely, with no inclination towards a particular probability.

Prior distributions on the other two parameters, that control the location and steepness of the S-curve, were chosen to provide very modest information to constrain estimation to the region of feasible values. Full details on prior selection are given in the supplementary appendix.

#### 2.3.2 Appraising model fits

Model-generated dose-event curves were inspected visually alongside source data to verify the quality of model fits. Furthermore, we recorded metrics for all Bayesian models that can diagnose a potentially poor model fit. Further details are given in the supplementary appendix.

#### 2.3.3 Software

Maximum-likelihood Emax models were fit using the DoseFinding package [145] and Bayesian models were fit using the brms package [146] in R [147]. Data processing was aided using tidyverse [148] packages, posterior samples were extracted using tidybayes [149], and plots were produced using ggplot2 [150].

## 3 Results

115 (94%) of the 122 examined manuscripts reported outcomes by dose-level. After extracting data and creating fully-orderable dose subsets, these yielded 155 analysis series for DLT, and 93 analysis series for OR (Figure 1). Characteristic information is summarised in Table 1.

**Table 1:** Characteristics of manuscripts studied and outcome series extracted.

|  | Manuscripts, N = 115 | DLT series, N = 155 | OR series, N = 93 |
| --- | --- | --- | --- |
| **Year of publication** |  |  |  |
| 2008 | 13 (11%) | 17 (11%) | 13 (14%) |
| 2009 | 9 (7.8%) | 14 (9.0%) | 10 (11%) |
| 2010 | 9 (7.8%) | 10 (6.5%) | 8 (8.6%) |
| 2011 | 20 (17%) | 23 (15%) | 11 (12%) |
| 2012 | 26 (23%) | 35 (23%) | 23 (25%) |
| 2013 | 16 (14%) | 22 (14%) | 16 (17%) |
| 2014 | 22 (19%) | 34 (22%) | 12 (13%) |
| **Experimental design** |  |  |  |
| Model-based | 85 (74%) | 116 (75%) | 56 (60%) |
| Rule-based | 30 (26%) | 39 (25%) | 37 (40%) |
| **Disease class** |  |  |  |
| Non-haematological | 82 (71%) | 117 (75%) | 71 (76%) |
| Haematological | 30 (26%) | 31 (20%) | 18 (19%) |
| Both | 2 (1.7%) | 6 (3.9%) | 4 (4.3%) |
| Not disclosed | 1 (0.9%) | 1 (0.6%) | 0 (0%) |
| **Disease** |  |  |  |
| Solid tumours |  | 48 (31%) | 29 (31%) |
| Breast cancer |  | 9 (5.8%) | 8 (8.6%) |
| Gastrointestinal cancer |  | 10 (6.5%) | 6 (6.5%) |
| AML |  | 7 (4.5%) | 6 (6.5%) |
| Lung cancer |  | 9 (5.8%) | 3 (3.2%) |
| Lymphoma |  | 6 (3.9%) | 5 (5.4%) |
| Multiple myeloma |  | 6 (3.9%) | 4 (4.3%) |
| Glioma |  | 5 (3.2%) | 4 (4.3%) |
| Melanoma |  | 4 (2.6%) | 4 (4.3%) |
| Mixed haematological cancers |  | 8 (5.2%) | 0 (0%) |
| CNS tumours |  | 4 (2.6%) | 3 (3.2%) |
| Head and neck cancer |  | 4 (2.6%) | 3 (3.2%) |
| Brain cancer |  | 5 (3.2%) | 1 (1.1%) |
| Lymphoma and advanced solid tumours |  | 4 (2.6%) | 2 (2.2%) |
| Sarcoma |  | 3 (1.9%) | 3 (3.2%) |
| CLL |  | 3 (1.9%) | 2 (2.2%) |
| Hepatocellular carcinoma |  | 2 (1.3%) | 2 (2.2%) |
| Renal cell carcinoma |  | 2 (1.3%) | 2 (2.2%) |
| Biliary tract cancer |  | 3 (1.9%) | 0 (0%) |
| Glioblastoma |  | 2 (1.3%) | 1 (1.1%) |
| NSCLC |  | 2 (1.3%) | 1 (1.1%) |
| SCLC |  | 3 (1.9%) | 0 (0%) |
| Cervical cancer |  | 1 (0.6%) | 1 (1.1%) |
| Colorectal cancer |  | 1 (0.6%) | 1 (1.1%) |
| Gynaecological cancer |  | 1 (0.6%) | 1 (1.1%) |
| B-cell malignancies |  | 0 (0%) | 1 (1.1%) |
| Not disclosed |  | 1 (0.6%) | 0 (0%) |
| Pancreatic cancer |  | 1 (0.6%) | 0 (0%) |
| Rectal cancer |  | 1 (0.6%) | 0 (0%) |
| **Treatment undergoing dose-escalation** |  |  |  |
| Inhibitor |  | 58 (37%) | 43 (46%) |
| Chemotherapy |  | 51 (33%) | 25 (27%) |
| Chemotherapy + inhibitor |  | 17 (11%) | 12 (13%) |
| Monoclonal antibody |  | 9 (5.8%) | 1 (1.1%) |
| Immunomodulatory |  | 4 (2.6%) | 2 (2.2%) |
| Immunomodulatory + chemotherapy |  | 3 (1.9%) | 2 (2.2%) |
| Radiotherapy |  | 4 (2.6%) | 1 (1.1%) |
| Oncolytic virus |  | 2 (1.3%) | 2 (2.2%) |
| Radiopharmaceutical + inhibitor |  | 2 (1.3%) | 2 (2.2%) |
| Cell therapy |  | 1 (0.6%) | 2 (2.2%) |
| Cytokine |  | 1 (0.6%) | 1 (1.1%) |
| Chemoprevention |  | 1 (0.6%) | 0 (0%) |
| Gene therapy |  | 1 (0.6%) | 0 (0%) |
| Not disclosed |  | 1 (0.6%) | 0 (0%) |
| Treatment ensemble contains chemotherapy |  | 91 (59%) | 49 (53%) |
| Number of dose-levels |  | 4 (2, 5) | 4 (2, 5) |

**Figure 1:**
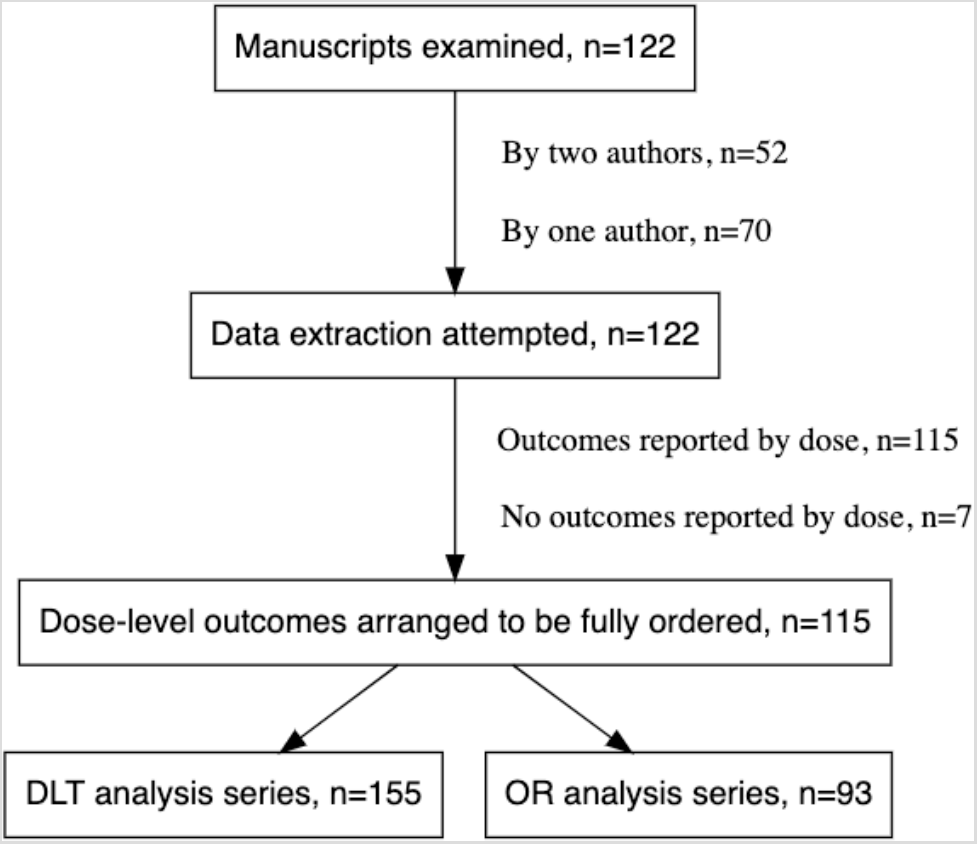
The data extraction process. The procedure for forming fully orderable subsets of doses is described in the text.

### 3.1 Patient groups

Three-quarters of the data analysed came from non-haematological cancers. In approximately one third of cases, the patient group comprised different types of solid tumour. Most commonly, however, trials were conducted within specific cancer sites, with breast, gastrointestinal, and lung cancers featuring relatively frequently.

Approximately one quarter of the data came from trials in haematological malignancies. Once again, trials were sometimes conducted in fairly heterogeneous patient groups and sometimes in specific diseases like AML, CLL and lymphoma.

### 3.2 Experimental treatments

The treatment type most commonly undergoing dose-escalation was inhibitors, contributing 58 (37%) DLT series and 43 (46%) OR series. Chemotherapies were also common targets for dose-escalation, contributing 51 (33%) DLT series and 25 (27%) OR series. Monoclonal antibodies were fairly infrequent in this data set, contributing only 9 (6%) DLT series and 1 (1%) OR series. Trials that escalated dose in two different treatment types were common, with chemotherapy plus inhibitor the most common pairing, yielding 11% of the DLT series and 13% of the OR series. The median of doses in an analysis series was 4 (IQR = 2, 5).

### 3.3 Monotonicity of DLT and OR in dose

Fitted curves for the dose-DLT series produced by the Bayesian Emax models are shown in Figure 2. Each line reflects the best fit to all of the DLT outcomes observed in one analysis series. A separate panel is shown for each type of treatment that underwent dose-escalation. Information on patient group and concomitant therapies are not shown in this plot.

**Figure 2:**
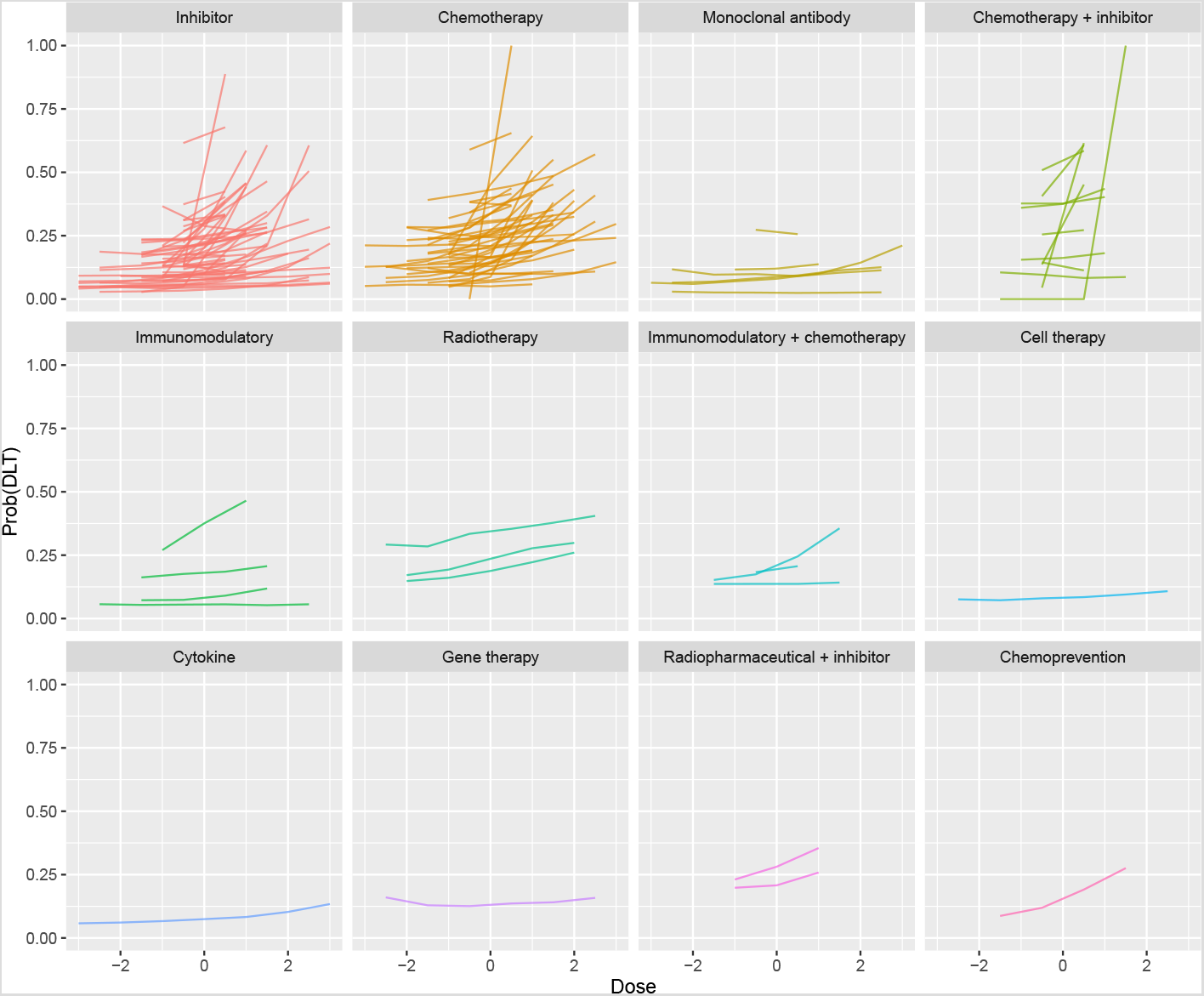
Fitted dose-DLT curves yielded by Bayesian Emax models. For presentation, doses are centralised at zero (i.e. the average dose-level for each series is subtracted) to allow the series to be visualised together.

We see that the majority of the fitted DLT series show a positive relationship with dose, reflecting that DLT becomes more likely as dose is increased. This is seen in all types of therapy and matches our expectation that the dose makes the poison. The fitted series for inhibitors and chemotherapies appear to increase more steeply than for other therapies.

Fitted curves for the dose-OR series produced by the Bayesian Emax models are shown in Figure 3. Contrasting to Figure 2, we see that there are materially fewer OR curves than DLT curves. However, it is clear that the fitted OR curves are much less likely to show a strong positive association between dose and response. It is striking how few positive gradients there are. Even amongst chemotherapies, there is scant evidence of greater efficacy at higher doses. The single OR series for a monoclonal antibody and one of the series for an immunomodulatory drug show comparatively strong evidence of a positive dose-response effect.

**Figure 3:**
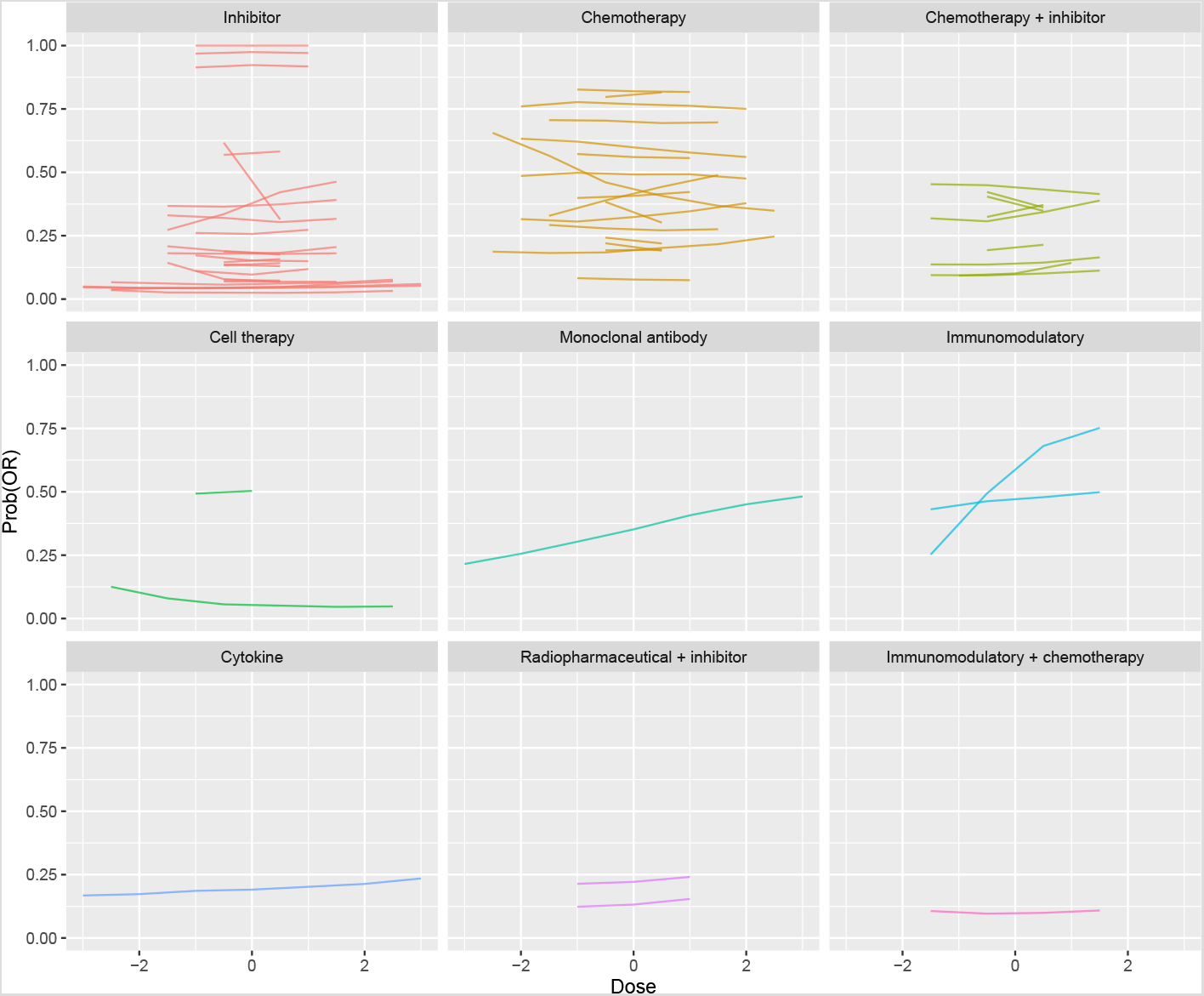
Fitted dose-OR curves yielded by Bayesian Emax models. For presentation, doses are centralised at zero (i.e. the average dose-level for each series is subtracted) to allow the series to be visualised together.

The heights of the fitted dose-DLT and dose-OR curves are shown in Figure 4, with statistics for the two outcomes plotted side-by-side within treatment type. The dashed red line at zero reflects the null value where there is no relationship between dose and event. Positive curve heights indicate that the event became more likely as dose is increased, and vice-versa.

**Figure 4:**
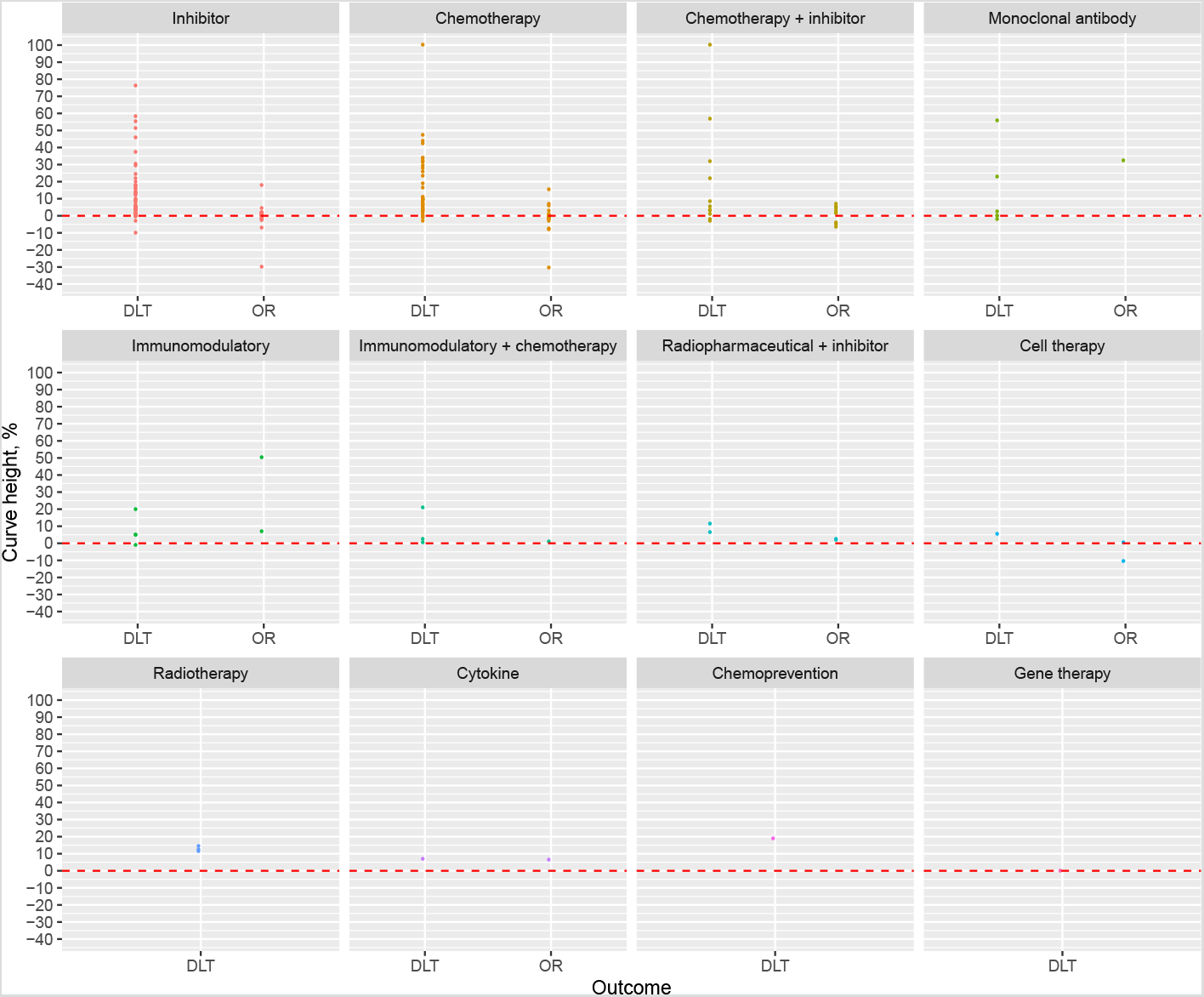
Heights of dose-DLT and dose-OR curves. The dashed red lines reflect a curve height of zero where there is no association between dose and event. Positive values reflect that event probabilities increase as dose is increased, and negative values reflect a decreasing probability as dose is increased.

**Figure 5:**
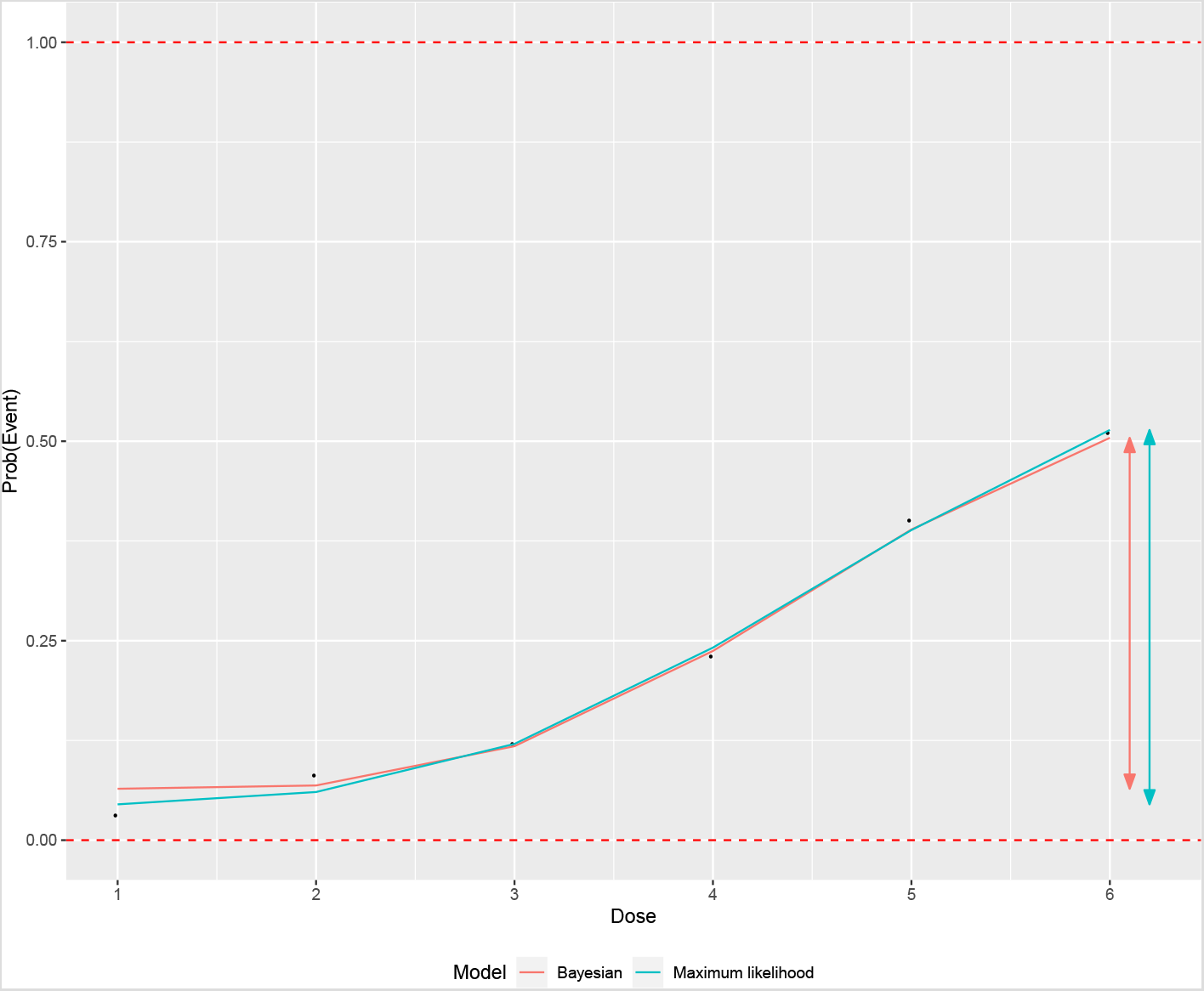
Visual demonstration of fitted Emax curves and the concept of curve height. *n* = 100 binary event outcomes were simulated at each of six dose-levels with event probabilities (0.01, 0.07, 0.12, 0.25, 0.47, 0.52). The heights of the fitted curves are demonstrated by the vertical lines with arrows on the right-hand side, measuring the distance from the fitted curve at the lowest dose to the highest dose. A modest amount of difference is visible between the lines fit by maximum likelihood and Bayesian models.

We see that the majority of DLT curves show a positive curve height in all classes of treatment. There are infrequent series that suggest no relationship, or a modestly negative relationship. In stark contrast, the same statistics for the OR series straddle the null line in the most frequent treatment categories, demonstrating a lack of evidence in support of the monotonic efficacy assumption.

Furthermore, the curve heights of the DLT series show much greater variability than those of the OR series. There are some instances where the probability of DLT increased quite rapidly as dose increased.

Curve heights for DLT and OR series are plotted by type of disease in Figure 7 in the supplementary appendix. We see that the phenomena we have described are broadly observed across most disease types. Figure 8 in the supplementary appendix shows the equivalent inferences split by class of dose-finding methodology (i.e. rule-based or model-based). The two OR series with the steepest positive relationship with dose both use a rule-based design. Generally, however, the heights of the DLT and OR curves from studies that used model-based designs were similar to those yielded by rule-based designs. In both groups, the observation remained that DLT curves commonly increased with dose whilst OR curves were mostly invariant in dose.

**Figure 6:**
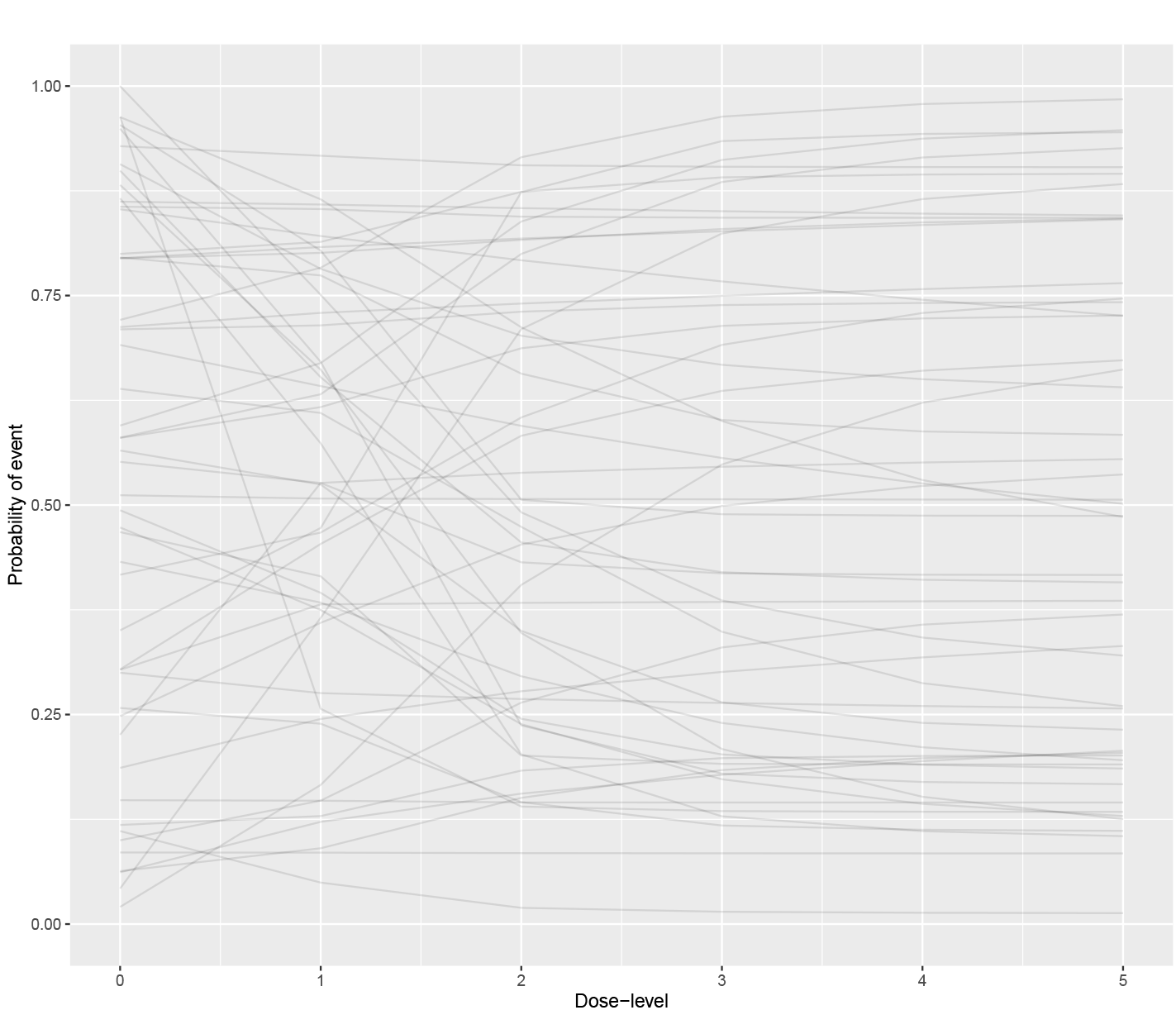
Prior predictive dose-event curves. A candidate value for each of the four Emax parameters was sampled from each of the described prior distributions. The fitted values determined by these parameter values were then calculated using the Emax model with *D* = 0, 1, 2, 3, 4, 5 to produce a single fitted curve. This process was then repeated fifty times. The same priors were used for both DLT and OR outcomes.

**Figure 7:**
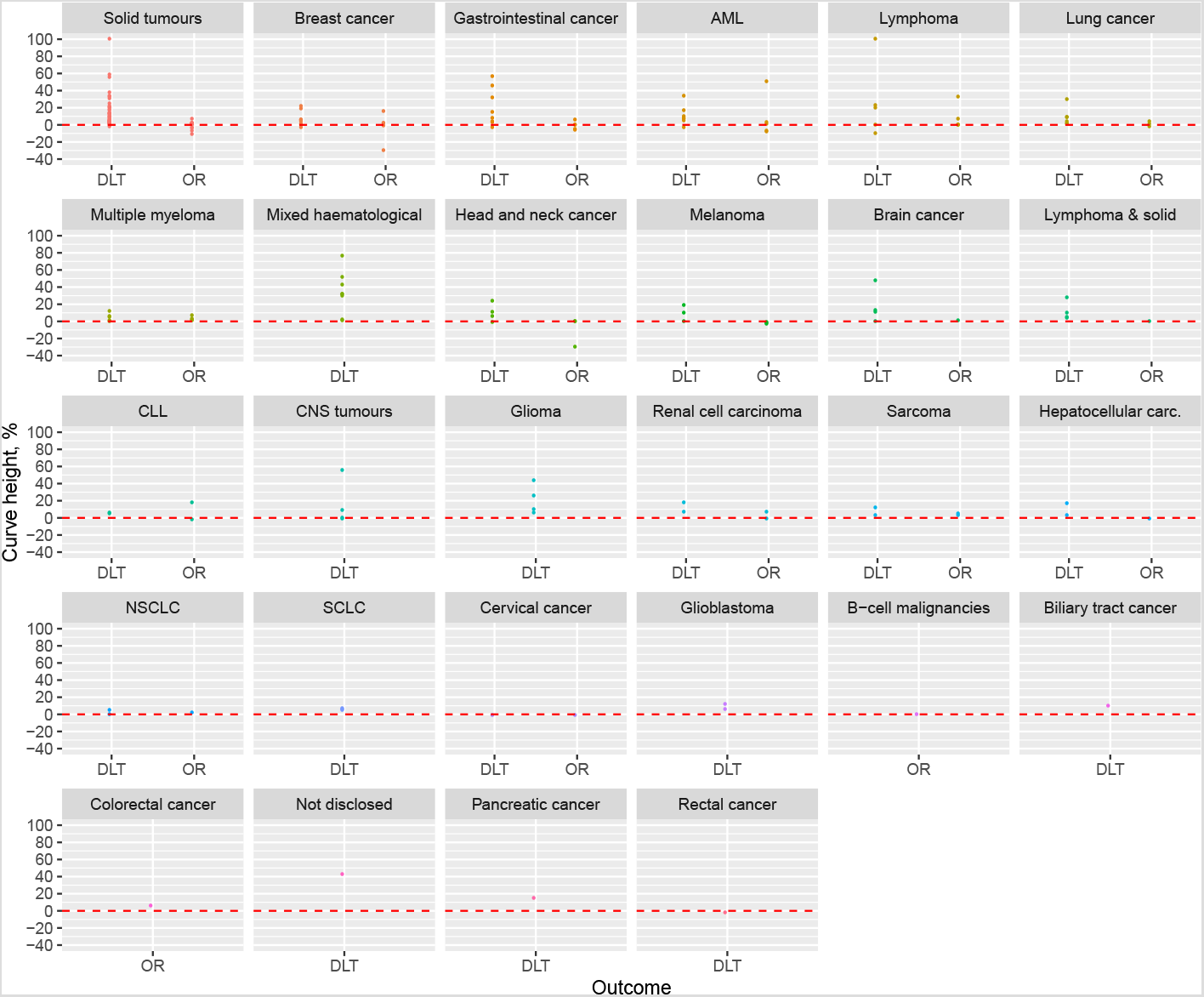
Heights of dose-DLT and dose-OR curves by type of disease. The dashed red lines reflect a curve height of zero where there is no association between dose and event. Positive values reflect that event probabilities increase as dose is increased, and negative values reflect a decreasing probability as dose is increased.

**Figure 8:**
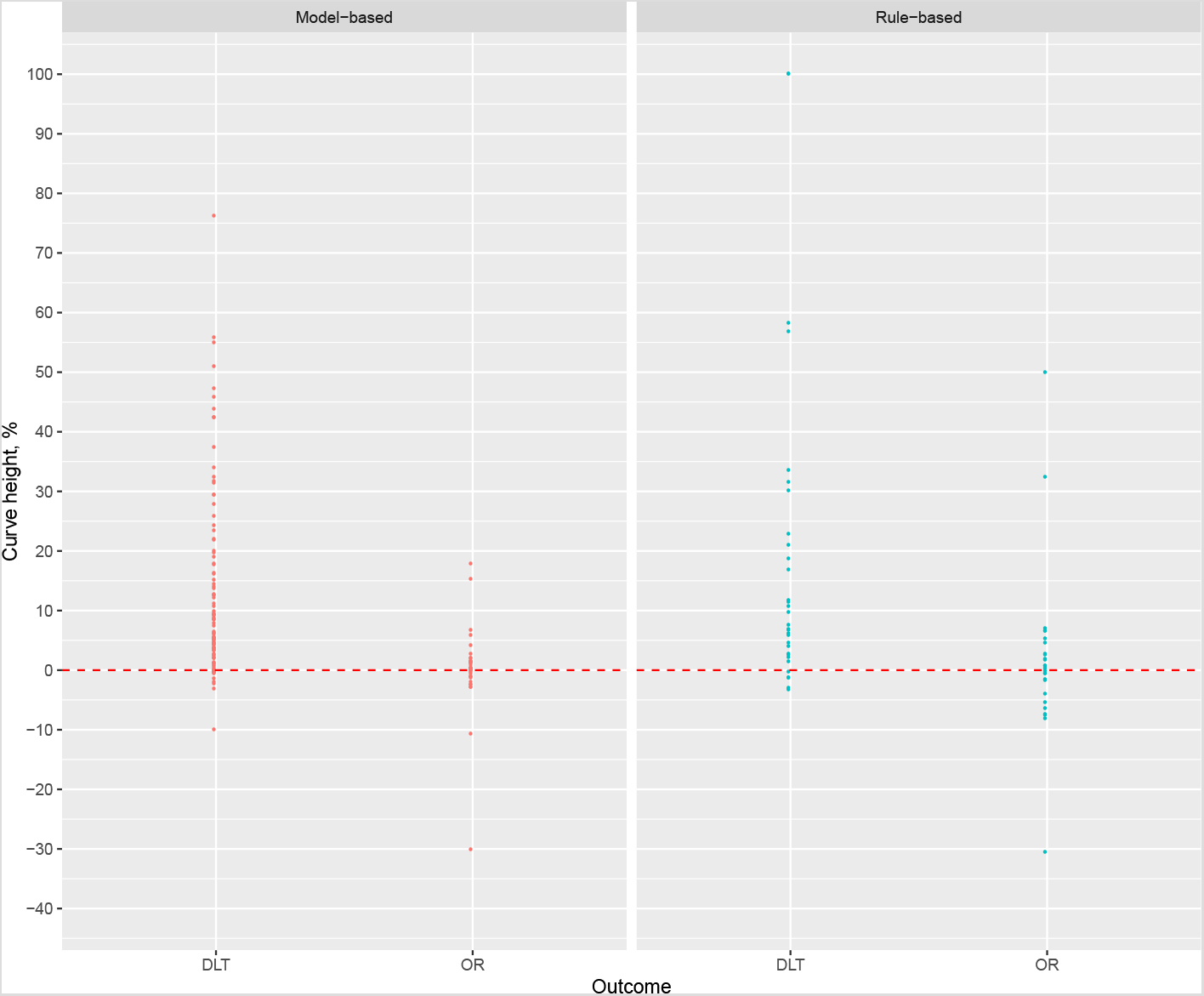
Heights of dose-DLT and dose-OR curves by class of experimental dose-finding design. The dashed red lines reflect a curve height of zero where there is no association between dose and event. Positive values reflect that event probabilities increase as dose is increased, and negative values reflect a decreasing probability as dose is increased.

The inferences thus far have come from the Bayesian Emax models, where the statistical model fitting procedure succeeded in all instances. We also sought to analyse series using maximum likelihood Emax models. The maximum likelihood model fitting procedure failed in many instances. Examples of model-fitting failure were procedures that did not converge, or procedures that yielded extremely large estimates for the standard errors of model parameters. For completeness, we show the fitted DLT and OR series yielded by the maximum likelihood models in Figures 9 and 10 in the supplementary appendix. The fitted series produced by the maximum likelihood models are less smooth. Figure 11 shows that the curve heights estimated by maximum likelihood models are generally more extreme than those produced by Bayesian models, with more values positive and negative values far from zero for DLT and OR series. In the most frequently investigated treatments, once again we see that the DLT series are overwhelmingly likely to have a positive curve height, whilst the heights of OR series cluster around zero.

**Figure 9:**
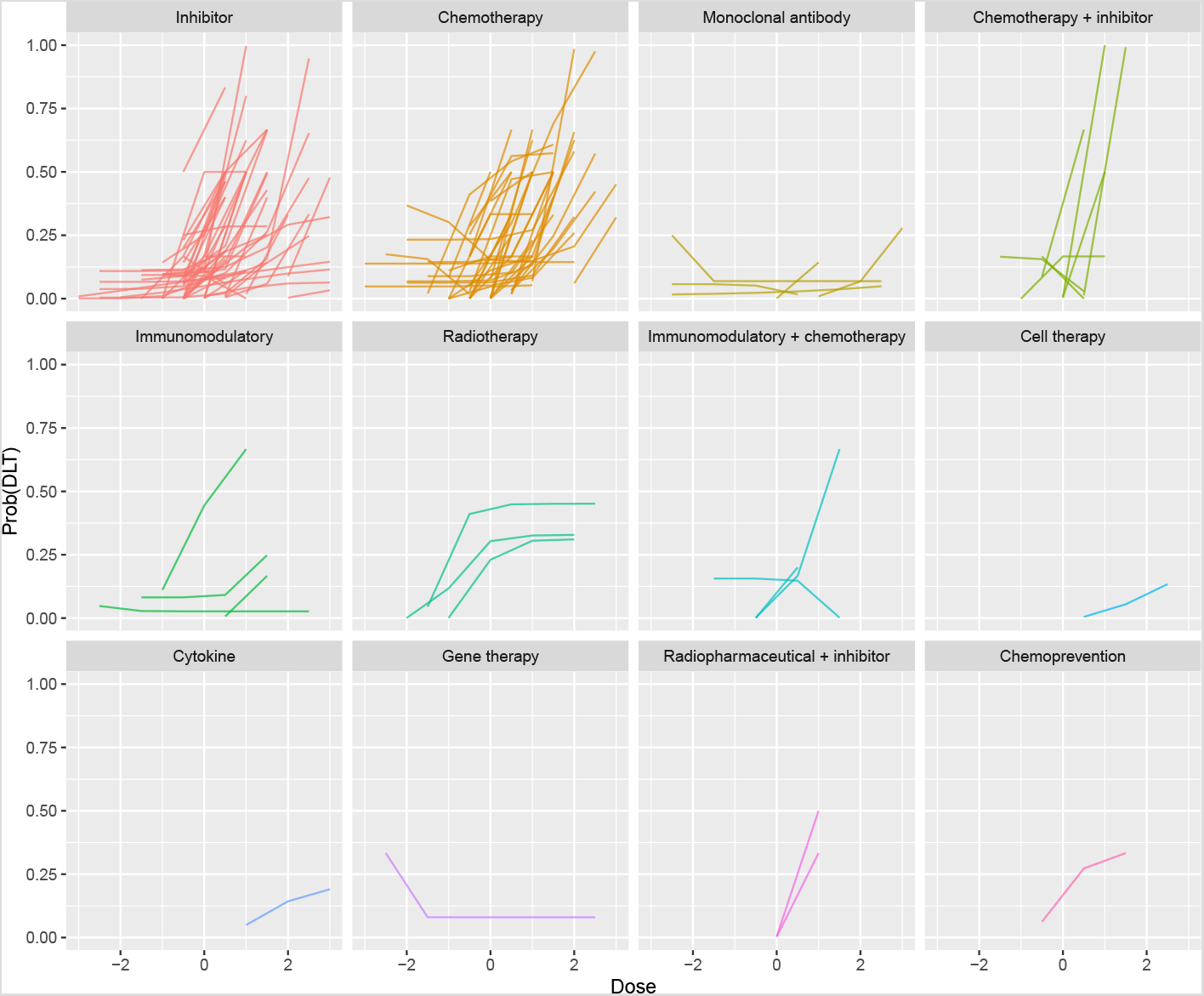
Fitted dose-DLT curves yielded by maximum likelihood models. For presentation, doses are centralised at zero (i.e. the average dose-level for each series is subtracted) to allow the series to be visualised together.

**Figure 10:**
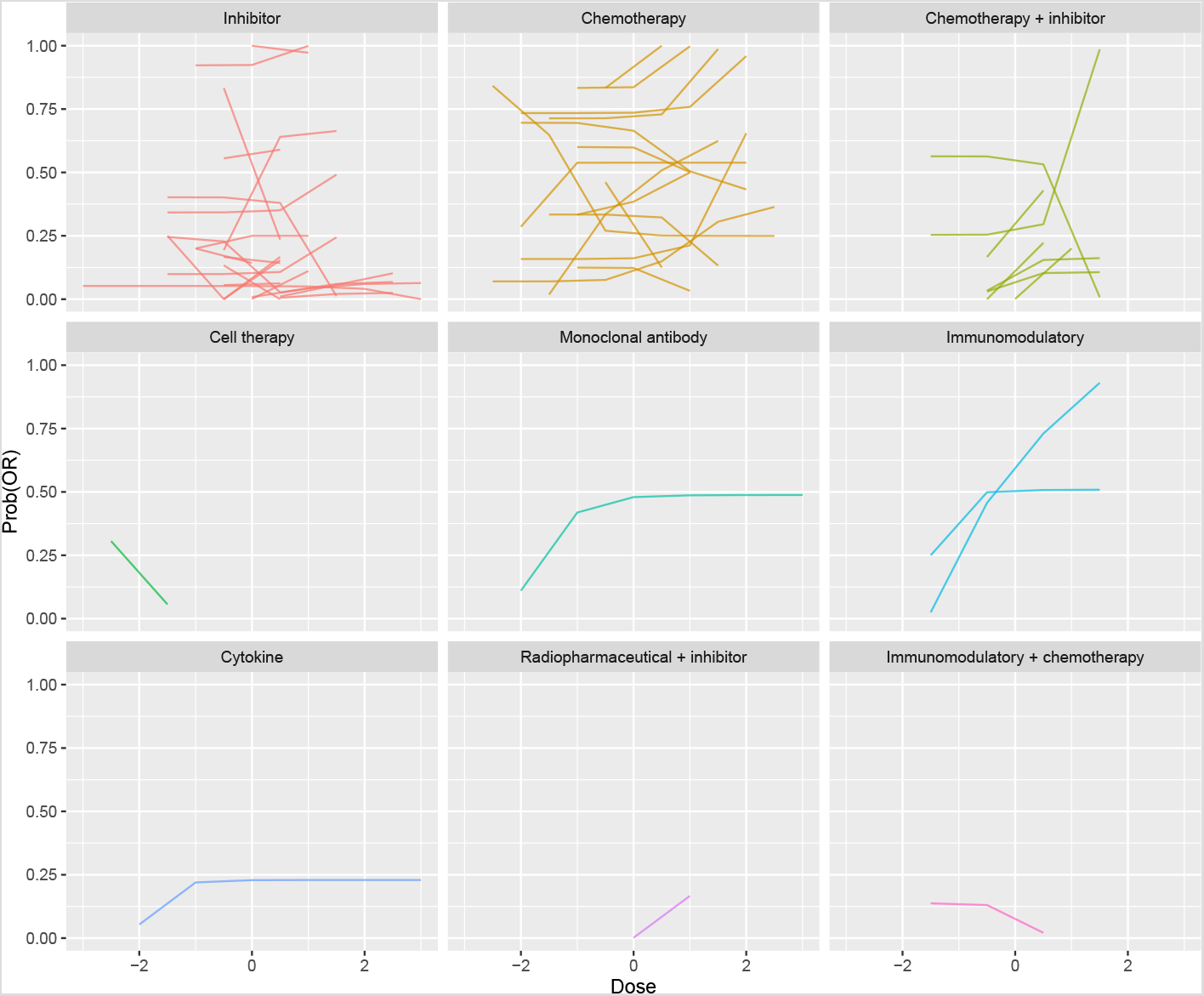
Fitted dose-OR curves yielded by maximum likelihood models. For presentation, doses are centralised at zero (i.e. the average dose-level for each series is subtracted) to allow the series to be visualised together.

**Figure 11:**
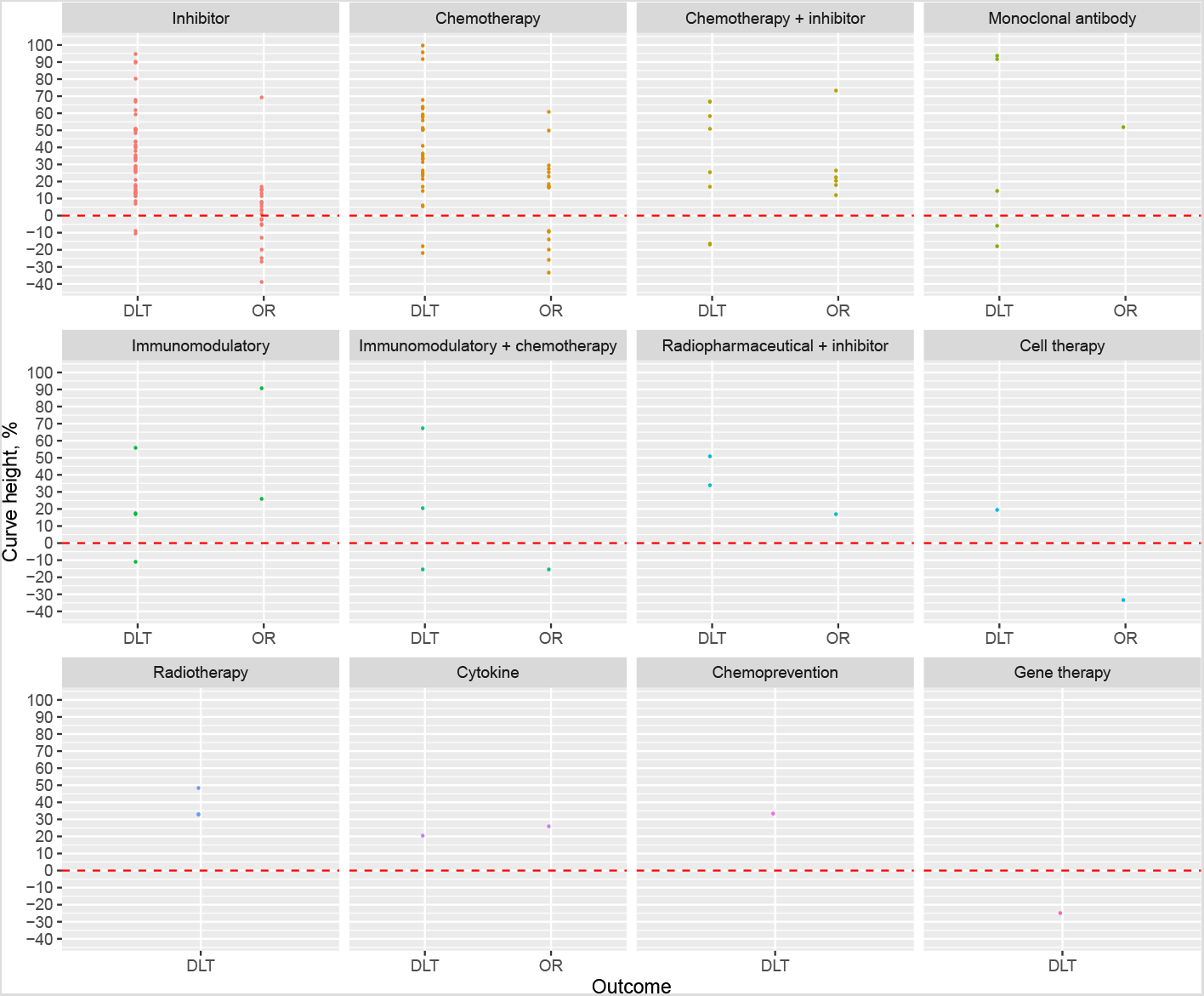
Heights of dose-DLT and dose-OR curves estimated by maximum-likelihood models. The dashed red lines reflect a curve height of zero where there is no association between dose and event. Positive values reflect that event probabilities increase as dose is increased, and negative values reflect a decreasing probability as dose is increased.

## 4 Discussion

We collected dose-level toxicity and efficacy outcomes from 115 papers reporting early phase clinical trials of putative anti-cancer therapies. These trials all used experimental methods that assume that higher doses are associated with greater probabilities of both toxicity and efficacy. We then analysed those outcomes using flexible non-linear regression methods. In summary, we found broad evidence that the probability of toxicity increased in dose in most treatment classes, in most types of cancer, in scenarios that use rule-based or model-based dose-escalation methodologies. In contrast, we found very little evidence that the probability of response increased as dose was increased.

On the face of it, the implication of our findings is that dose-escalation clinical trials commonly advocate doses that are unjustifiably high. For a treatment where toxicity incidence is positively associated with dose and response incidence is invariant to dose, lower doses should be preferred. However, by conducting dose-escalation experiments that recommend doses only by toxicity outcomes, explicitly assuming that higher doses are more efficacious and therefore preferable, many trials miss the opportunity to recommend a lower dose with less toxic effects and no commensurate cost to efficacy. This could help explain why dose-reduction occurs in phase II and III clinical trials. For example, a detailed account of post-phase I dose optimisation in an inhibitor drug is given by Lee *et al*. [151].

The statements above pertain to the ranges of doses investigated in phase I clinical trials. They do not apply to the ranges of all possible doses. Naturally, we acknowledge in all active therapies that there must be a dose so low that the attendant anti-tumour effect is negligible. That we found many series with non-trivial response rates at all doses is perhaps testament to the value of preclinical research and PK/PD modelling in locating dose ranges that are likely to be tolerable and active for phase I trials.

A logical remedy to the problems we have described would be to use so-called seamless phase I/II designs that recommend doses by toxicity and response outcomes, several examples of which have been published [9–13] and implemented in clinical trials [14, 152]. These designs bring their own challenges, the most notable of which are the extra statistical complexity and the requirement that the co-primary outcomes can be evaluated over a similar time horizon to allow dose-escalation decisions to be made in a timely manner.

A simpler solution would require early phase trialists to address the assumptions made by their phase I designs and discuss the appropriateness of their final dose recommendations in light of toxicity and efficacy outcomes. By reporting suitable efficacy and toxicity outcomes by dose, researchers would allow the community to assess the suitability of the methodological assumptions and identify doses most appropriate for further study. Arguably, this already occurs in practice when trialists use dose-escalation methods like 3+3, CRM or EWOC cognisant of the possibility that higher doses might not bring greater efficacy. The drawback of this putative approach is that it risks allocating patients in the dose-finding trial to inappropriate doses. Trialists that regard this as unethical would be encouraged to use seamless phase I/II methodologies, described above.

It is possible that monotonic efficacy effects in dose are observed in long-term clinical outcomes like OS and PFS, even if they are not seen in surrogate outcomes like response. We cannot provide any evidence to support or refute this hypothesis since survival outcomes were reported by dose extremely infrequently. If this hypothesis is true, however, it calls into question the validity of objective response as a surrogate outcome for clinical outcomes.

The sample size of the trials included in this research is small. Researchers might legitimately ponder the feasibility of detecting strong dose-event relationships with such small sample sizes. It was advantageous, then, that we included and analysed DLT outcomes because they have shown that evidence of stark relationships can be garnered from small trials, particularly when analysed together. Within study, OR outcomes routinely failed to show the strength of relationship with dose shown by DLT outcomes.

It is perhaps self-fulfilling that we have observed stark monotonic effects in toxicity because dose-finding trials escalate dose in the absence of unacceptable toxicity and de-escalate dose or halt the trial when toxicity manifests. For this reason, it is plausible that the toxicity relationships we have presented here may be biased upwards. Nevertheless, this does not explain the lack of evidence for a relationship between dose and efficacy.

Recently, Hazim *et al*. [153] investigated the relationship between dose and efficacy in single-agent phase I trials in oncology. They did so by calculating aggregate response rates at dose categories defined by the ratio of the given dose to the recommended phase II dose (RP2D). They found evidence that response rates increased modestly as doses approached the RP2D. Compared to their study, this research benefits from including treatment combinations, including both efficacy and toxicity outcomes, and using statistical modelling.

We conclude this article with some advice to investigators on the design, conduct and reporting of dose-escalation clinical trials. If response is a validate surrogate for efficacy in your patient group, consider using co-primary toxicity and response outcomes in a phase I/II dose-finding design [9–13]. If you are doubtful of the validity of response as a surrogate in your patient group, consider how you will assess whether a higher dose is associated with better outcomes for patients. When selecting a design, be aware of the assumptions of the methodologies under consideration. If you use a method that assumes higher doses are better, justify why this is appropriate. When reporting your trial, clearly state your measures of toxicity and efficacy and report outcomes broken down by dose. Be mindful of the assumptions inherent in your experimental design, how they agree or disagree with the observed outcomes, and the ramifications for patients via the doses recommended. Resist the temptation to pool outcomes across doses. Remember that the goal of dose-finding trials is to evaluate outcomes under a set of investigational doses and pooling outcomes obfuscates that goal.

## 5 End matter

### 5.1 Author contributions

KB wrote the manuscript. KB, VH and GS analysed the data. All authors contributed data. All authors contributed to the design of study and interpretation of results. All authors reviewed the manuscript and agreed to its submission.

### 5.2 Availability of materials

The data set analysed in this manuscript is available from a repository [143] hosted by the University of Birmingham. R code to reproduce all statistics, tables and figures is freely available at https://github.com/brockk/dosefindingdata.

## Data Availability

All data and materials are available from public repositories. URLS are given below.

https://github.com/brockk/dosefindingdata

https://doi.org/10.25500/edata.bham.00000337

## 6 Supplementary Appendix

### 6.1 Supplementary Methods

#### 6.1.1 Graphical illustration of curve height

#### 6.1.2 Prior distributions

The canonical form for the Emax model is

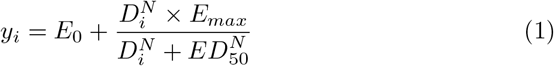

where

1. *y_i_* is the event probability under the dose given to patient *i*;
2. *D_i_* is the dose given to patient *i*;
3. *E*_0_ is the event probability when exposure is zero, or the zero-dose effect;
4. *E_max_* is the maximum effect attributable to dosing, i.e. the height of the sigmoidal curve;
5. *ED*_50_ is the dose that produces half of *E_max_*;
6. *N >* 0 is the slope factor, determining the steepness of the dose-event curve.

This parameterisation, however, presents difficulties when specifying priors for a binary response. For a binary outcome, the response variable is a probability and is thus defined only on the closed interval [0, 1]. Under the parameterisation above, we require that *E*_0_ *∈* [0, 1] and *E*_0_ + *E_max_ ∈* [0, 1]. However, it is difficult to specify a prior on the sum of two parameters. It is much easier to respecify the Emax model as:

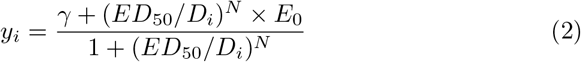

where

1. *γ* = *E*_0_ + *E_max_* is the maximum event probability;
2. *•* and all other variables retain their previous interpretation.

It is now simple to ensure that *E*_0_ *∈* [0, 1] and *γ ∈* [0, 1] using priors. Our research question involves investigating the values estimated for *E*_0_ and *γ* in different clinical trials. The event rates that manifest depend on the activity of the treatment and doses selected for investigation. A completely inert treatment could produce event rates of zero. Excessively low doses of a treatment that otherwise has the potential to be active could also produce event rates of zero. Likewise, particularly active or toxic therapies could produce very high event rates, even at the lower doses of those under investigation. Each of these phenomena is plausible for therapies at the beginning of their clinical research pathway like those investigated in dose-finding trials. Thus, it is appropriate to place uniform priors on *E*_0_ and *γ* over the interval [0, 1].

We use folded-normal priors on *ED*_50_ and *N* with hard lower bounds at 0 because these parameters cannot take negative values, and no upper bounds.

In a study of *J* doses, we anticipate that *ED*_50_ takes the value *J/*2, i.e. we expect that the centre dose-level produces half of the maximum effect attributable to dosing. Naturally, however, we acknowledge that there is considerable uncertainty on the location of *ED*_50_ and set the prior standard deviation of *ED*_50_ to be *J −* 1. These hyperparameters ensure that the majority of the probability mass covers the doses under investigation whilst allocating approximately 25% of the mass to doses that exceed the maximum dose under investigation.

We use a folded-normal prior on *N* with hard lower bound at 0 and no upper bound. Greater values for *N* reflect steeper dose-event curves. The value *N* = 1 corresponds to a special null case of the Emax model called the hyperbolic model [144]. We assume a prior mean of 1 for *N*. Again, we acknowledge the material uncertainty in this variable, selecting a prior standard deviation of 3. After allowing for the lower bound, this yields a 90% prior credible interval for *N* of(0.25, 6.56).

Figure 6 shows 50 dose-event curves sampled from the prior predictive distribution in a scenario with five doses. These were constructed by sampling values for the parameters from the prior distributions described above and then plugging the values into (2) with *D_i_* = 0, 1, …, 5. In this manner, a single set of parameter values produces one dose-event curve. The process was then repeated 50 times.

In this modestly-sized sample, we see dose-event curves that start low and then increase, as we might expect to uncover for dose-toxicity curves in cytotoxic treatments. We see curves that start low and stay low, that could be consistent with inert treatments. We also see curves that start high and stay high, consistent with treatments where all the investigative doses are in the active or toxic range. Finally, we see some curves that start high and then decrease. We might not expect to see many of these in reality but it is preferable that their existence is admitted by our parameter prior choices. Furthermore, curves can plateau at any event probability between 0 and 1. Our prior parameters generate data that are consistent with our broad range of expectations of dose-event curves.

#### 6.1.3 Appraising Bayesian model fits

For all Bayesian models, we recorded the number of divergent transitions, the number of times the maximum tree-depth was exceeded, and the number of Monte Carlo chains with low Bayesian fraction of missing information (BFMI). The presence of each of these features can signal poor quality sampling that does not reflect the true posterior distribution.

### 6.2 Supplementary Results

#### 6.2.1 Curve heights by disease

#### 6.2.2 Curve heights by experimental design

#### 6.2.3 Fitted series and curve heights by maximum likelihood models

#### 6.2.4 Bayesian model fit diagnostics

None of the fitted Bayesian model fits exceeded the maximum tree-depth in sampling, or suffered from low BFMI. Seven of the DLT models and seven of the OR models had at least one divergent transition amongst the 4000 samples sought from the joint posterior distribution in each instance. Twelve of these fourteen had fewer than seven divergent transitions. Inferences from all fourteen instances were checked by plotting the sampled dose-event curves alongside the observed trial outcomes to ensure that the posterior inferences were a faithful representation of the data in each case.

## Notes

### Competing Interest Statement

The authors have declared no competing interest.

### Funding Statement

Kristian Brock is funded by the University of Birmingham.

### Author Declarations

This research collates previously published data and requires no IRB / oversight approval.

## References

1 A. Rogatko, D. Schoeneck, W. Jonas, M. Tighiouart, F. R. Khuri, and A. Porter, “Translation of innovative designs into phase I trials,” Journal of Clinical Oncology, vol. 25, no. 31, pp. 4982–4986, 2007. ISBN: 1527-7755 (Electronic) 0732-183X (Linking).

2 C. Chiuzan, J. Shtaynberger, G. A. Manji, J. K. Duong, G. K. Schwartz, A. Ivanova, and S. M. Lee, “Dose-finding designs for trials of molecularly targeted agents and immunotherapies,” Journal of Biopharmaceutical Statistics, vol. 27, no. 3, pp. 477–494, 2017. Publisher: Taylor & Francis.

3 C. Le Tourneau, J. J. Lee, and L. L. Siu, “Dose escalation methods in phase i cancer clinical trials,” Journal of the National Cancer Institute, vol. 101, no. 10, pp. 708–720, 2009.

4 J. O’Quigley, M. Pepe, and L. Fisher, “Continual reassessment method: A practical design for phase 1 clinical trials in cancer.,” Biometrics, vol. 46, no. 1, pp. 33–48, 1990.

5 M. Tighiouart and A. Rogatko, “Dose Finding with Escalation with Overdose Control (EWOC) in Cancer Clinical Trials,” Statistical Science, vol. 25, no. 2, pp. 217–226, 2010. arXiv: 1011.6479v1.

6 J. O’Quigley and S. Zohar, “Experimental designs for phase i and phase i/ii dose-finding studies,” British journal of cancer, vol. 94, no. 5, pp. 609–613, 2006.

7 A. Iasonos, A. S. Wilton, E. R. Riedel, V. E. Seshan, and D. R. Spriggs, “A comprehensive comparison of the continual reassessment method to the standard 3+ 3 dose escalation scheme in phase i dose-finding studies,” Clinical Trials, vol. 5, no. 5, pp. 465–477, 2008.

8 G. M. Wheeler, A. P. Mander, A. Bedding, K. Brock, V. Cornelius, A. P. Grieve, T. Jaki, S. B. Love, C. J. Weir, C. Yap, et al., “How to design a dose-finding study using the continual reassessment method,” BMC medical research methodology, vol. 19, no. 1, pp. 1–15, 2019.

9 T. M. Braun, “The bivariate continual reassessment method: Extending the CRM to phase I trials of two competing outcomes,” Controlled Clinical Trials, vol. 23, no. 3, pp. 240–256, 2002.

10 W. Zhang, D. J. Sargent, and S. Mandrekar, “An adaptive dose-finding design incorporating both toxicity and efficacy,” Statistics in Medicine, vol. 25, no. 14, pp. 2365–2383, 2006.

11 A. Ivanova, “A new dose-finding design for bivariate outcomes,” Biometrics, vol. 59, no. 4, pp. 1001–1007, 2003.

12 P. Thall and J. Cook, “Dose-Finding Based on Efficacy-Toxicity Trade-Offs,” Biometrics, vol. 60, no. 3, pp. 684–693, 2004.

13 N. A. Wages and C. Tait, “Seamless phase i/ii adaptive design for oncology trials of molecularly targeted agents,” Journal of biopharmaceutical statistics, vol. 25, no. 5, pp. 903–920, 2015.

14 K. Brock, L. Billingham, M. Copland, S. Siddique, M. Sirovica, and C. Yap, “Implementing the efftox dose-finding design in the matchpoint trial,” BMC medical research methodology, vol. 17, no. 1, p. 112, 2017.

15 A. Patnaik, S. P. Kang, D. Rasco, K. P. Papadopoulos, J. Elassaiss-Schaap, M. Beeram, R. Drengler, C. Chen, L. Smith, G. Espino, et al., “Phase i study of pembrolizumab (mk-3475; anti–pd-1 monoclonal antibody) in patients with advanced solid tumors,” Clinical cancer research, vol. 21, no. 19, pp. 4286–4293, 2015.

16 E. B. Garon, N. A. Rizvi, R. Hui, N. Leighl, A. S. Balmanoukian, J. P. Eder, A. Patnaik, C. Aggarwal, M. Gubens, L. Horn, et al., “Pembrolizumab for the treatment of non–small-cell lung cancer,” New England Journal of Medicine, vol. 372, no. 21, pp. 2018–2028, 2015.

17 R. S. Herbst, P. Baas, D.-W. Kim, E. Felip, J. L. Pérez-Gracia, J.-Y. Han, J. Molina, J.-H. Kim, C. D. Arvis, M.-J. Ahn, et al., “Pembrolizumab versus docetaxel for previously treated, pd-l1-positive, advanced non-smallcell lung cancer (keynote-010): a randomised controlled trial,” The Lancet, vol. 387, no. 10027, pp. 1540–1550, 2016.

18 S. Sharma, E. G. de Vries, J. R. Infante, C. N. Oldenhuis, J. A. Gietema, L. Yang, S. Bilic, K. Parker, M. Goldbrunner, J. W. Scott, and H. A. Burris, “Safety, pharmacokinetics, and pharmacodynamics of the DR5 antibody LBY135 alone and in combination with capecitabine in patients with advanced solid tumors,” Investigational New Drugs, vol. 32, pp. 135–144, apr 2013.

19 A. Popovtzer, D. Normolle, F. P. Worden, M. E. Prince, D. B. Chepeha, G. T. Wolf, C. R. Bradford, T. S. Lawrence, and A. Eisbruch, “Phase i trial of radiotherapy concurrent with twice-weekly gemcitabine for head and neck cancer: Translation from preclinical investigations aiming to improve the therapeutic ratio,” Translational Oncology, vol. 7, pp. 479–483, aug 2014.

20 S. Kobayashi, H. Nagano, D. Sakai, H. Eguchi, E. Hatano, M. Kanai, S. Seo, K. Taura, Y. Fujiwara, T. Ajiki, S. Takemura, S. Kubo, H. Yanag-imoto, H. Toyokawa, A. Tsuji, H. Terajima, S. Morita, and T. Ioka, “Phase i study of adjuvant gemcitabine or s-1 in patients with biliary tract cancers undergoing major hepatectomy: KHBO1003 study,” Cancer Chemotherapy and Pharmacology, vol. 74, pp. 699–709, jul 2014.

21 I. F. Khouri, W. Wei, M. Korbling, F. Turturro, S. Ahmed, A. Alousi, P. Anderlini, S. Ciurea, E. Jabbour, B. Oran, U. R. Popat, G. Rondon, R. L. Bassett, and A. Gulbis, “BFR (bendamustine, fludarabine, and rituximab) allogeneic conditioning for chronic lymphocytic leukemia/lymphoma: reduced myelosuppression and GVHD,” Blood, vol. 124, pp. 2306–2312, oct 2014.

22 Y. Ando, M. Inada-Inoue, A. Mitsuma, T. Yoshino, A. Ohtsu, N. Suenaga, M. Sato, T. Kakizume, M. Robson, C. Quadt, and T. Doi, “Phase i dose-escalation study of buparlisib (BKM120), an oral pan-class i PI3k inhibitor, in japanese patients with advanced solid tumors,” Cancer Science, vol. 105, pp. 347–353, feb 2014.

23 A. T. Shaw, D.-W. Kim, R. Mehra, D. S. Tan, E. Felip, L. Q. Chow, D. R. Camidge, J. Vansteenkiste, S. Sharma, T. D. Pas, G. J. Riely, B. J. Solomon, J. Wolf, M. Thomas, M. Schuler, G. Liu, A. Santoro, Y. Y. Lau, M. Goldwasser, A. L. Boral, and J. A. Engelman, “Ceritinib in ALK-rearranged non–small-cell lung cancer,” New England Journal of Medicine, vol. 370, pp. 1189–1197, mar 2014.

24 C. Saura, J. Bendell, G. Jerusalem, S. Su, Q. Ru, S. D. Buck, D. Mills, S. Ruquet, A. Bosch, A. Urruticoechea, J. T. Beck, E. D. Tomaso, D. W. Sternberg, C. Massacesi, S. Hirawat, L. Dirix, and J. Baselga, “Phase ib study of buparlisib plus trastuzumab in patients with HER2-positive advanced or metastatic breast cancer that has progressed on trastuzumab-based therapy,” Clinical Cancer Research, vol. 20, pp. 1935–1945, jan 2014.

25 J. Rodon, H. A. Tawbi, A. L. Thomas, R. G. Stoller, C. P. Turtschi, J. Baselga, J. Sarantopoulos, D. Mahalingam, Y. Shou, M. A. Moles, L. Yang, C. Granvil, E. Hurh, K. L. Rose, D. D. Amakye, R. Dummer, and A. C. Mita, “A phase i, multicenter, open-label, first-in-human, dose-escalation study of the oral smoothened inhibitor sonidegib (LDE225) in patients with advanced solid tumors,” Clinical Cancer Research, vol. 20, pp. 1900–1909, feb 2014.

26 S. Faderl, K. Balakrishnan, D. A. Thomas, F. Ravandi, G. Borthakur, J. Burger, A. Ferrajoli, J. Cortes, S. O’Brien, T. Kadia, J. Feliu, W. Plunkett, V. Gandhi, and H. M. Kantarjian, “Phase i and extension study of clofarabine plus cyclophosphamide in patients with relapsed/refractory acute lymphoblastic leukemia,” Clinical Lymphoma Myeloma and Leukemia, vol. 14, pp. 231–238, jun 2014.

27 M. Boulin, P. Hillon, J. P. Cercueil, F. Bonnetain, S. Dabakuyo, A. Minello, J. L. Jouve, C. Lepage, M. Bardou, M. Wendremaire, P. Guerard, A. Denys, A. Grandvuillemin, B. Chauffert, L. Bedenne, and B. Guiu, “Idarubicin-loaded beads for chemoembolisation of hepatocellular carcinoma: results of the IDASPHERE phase i trial,” Alimentary Pharmacology & Therapeutics, vol. 39, pp. 1301–1313, apr 2014.

28 J. Rodon, I. Braña, L. L. Siu, M. J. D. Jonge, N. Homji, D. Mills, E. D. Tomaso, C. Sarr, L. Trandafir, C. Massacesi, F. Eskens, and J. C. Bendell, “Phase i dose-escalation and –expansion study of buparlisib (BKM120), an oral pan-class i PI3k inhibitor, in patients with advanced solid tumors,” Investigational New Drugs, vol. 32, pp. 670–681, mar 2014.

29 T. Doi, Y. Onozawa, N. Fuse, T. Yoshino, K. Yamazaki, J. Watanabe, M. Akimov, M. Robson, N. Boku, and A. Ohtsu, “Phase i dose-escalation study of the HSP90 inhibitor AUY922 in japanese patients with advanced solid tumors,” Cancer Chemotherapy and Pharmacology, vol. 74, pp. 629–636, jul 2014.

30 J. R. Infante, E. C. Dees, A. J. Olszanski, S. V. Dhuria, S. Sen, S. Cameron, and R. B. Cohen, “Phase i dose-escalation study of LCL161, an oral inhibitor of apoptosis proteins inhibitor, in patients with advanced solid tumors,” Journal of Clinical Oncology, vol. 32, pp. 3103–3110, oct 2014.

31 A. M. Tsimberidou, M. J. Keating, E. J. Jabbour, F. Ravandi-Kashani, S. O’Brien, E. Estey, N. Bekele, W. K. Plunkett, H. Kantarjian, and G. Borthakur, “A phase i study of fludarabine, cytarabine, and oxaliplatin therapy in patients with relapsed or refractory acute myeloid leukemia,” Clinical Lymphoma Myeloma and Leukemia, vol. 14, pp. 395–400.e1, oct 2014.

32 S. P. Iyer, J. T. Beck, A. K. Stewart, J. Shah, K. R. Kelly, R. Isaacs, S. Bilic, S. Sen, and N. C. Munshi, “A phase IB multicentre dose-determination study of BHQ880 in combination with anti-myeloma therapy and zoledronic acid in patients with relapsed or refractory multiple myeloma and prior skeletal-related events,” British Journal of Haematology, vol. 167, pp. 366–375, aug 2014.

33 I. Brana, R. Berger, T. Golan, P. Haluska, J. Edenfield, J. Fiorica, J. Stephenson, L. P. Martin, S. Westin, P. Hanjani, M. B. Jones, K. Almhanna, R. M. Wenham, D. M. Sullivan, W. S. Dalton, A. Gunchenko, J. D. Cheng, L. L. Siu, and J. E. Gray, “A parallel-arm phase i trial of the humanised anti-IGF-1r antibody dalotuzumab in combination with the AKT inhibitor MK-2206, the mTOR inhibitor ridaforolimus, or the NOTCH inhibitor MK-0752, in patients with advanced solid tumours,” British Journal of Cancer, vol. 111, pp. 1932–1944, oct 2014.

34 S. J. Isakoff, D. Wang, M. Campone, A. Calles, E. Leip, K. Turnbull, N. Bardy-Bouxin, L. Duvillié, and E. Calvo, “Bosutinib plus capecitabine for selected advanced solid tumours: results of a phase 1 dose-escalation study,” British Journal of Cancer, vol. 111, pp. 2058–2066, oct 2014.

35 R. C. Brennan, W. Furman, S. Mao, J. Wu, D. C. Turner, C. F. Stewart, V. Santana, and L. M. McGregor, “Phase i dose escalation and pharmacokinetic study of oral gefitinib and irinotecan in children with refractory solid tumors,” Cancer Chemotherapy and Pharmacology, vol. 74, pp. 1191–1198, sep 2014.

36 L. Gandhi, R. Bahleda, S. M. Tolaney, E. L. Kwak, J. M. Cleary, S. S. Pandya, A. Hollebecque, R. Abbas, R. Ananthakrishnan, A. Berkenblit, M. Krygowski, Y. Liang, K. W. Turnbull, G. I. Shapiro, and J.-C. Soria, “Phase i study of neratinib in combination with temsirolimus in patients with human epidermal growth factor receptor 2–dependent and other solid tumors,” Journal of Clinical Oncology, vol. 32, pp. 68–75, jan 2014.

37 M. Fanale, S. Assouline, J. Kuruvilla, P. Solal-Céligny, D. S. Heo, G. Verhoef, P. Corradini, J. S. Abramson, F. Offner, A. Engert, M. J. S. Dyer, D. Carreon, B. Ewald, J. Baeck, A. Younes, and A. S. Freedman, “Phase IA/II, multicentre, open-label study of the CD40 antagonistic monoclonal antibody lucatumumab in adult patients with advanced non-hodgkin or hodgkin lymphoma,” British Journal of Haematology, vol. 164, pp. 258–265, nov 2013.

38 P. Das, C. Eng, M. A. Rodriguez-Bigas, G. J. Chang, J. M. Skibber, Y. N. You, D. M. Maru, M. F. Munsell, M. V. Clemons, S. E. Kopetz, C. R. Garrett, I. Shureiqi, M. E. Delclos, S. Krishnan, and C. H. Crane, “Preoperative radiation therapy with concurrent capecitabine, bevacizumab, and erlotinib for rectal cancer: A phase 1 trial,” International Journal of Radiation Oncology Biology Physics, vol. 88, pp. 301–305, feb 2014.

39 B. Besse, R. Heist, V. Papadmitrakopoulou, D. Camidge, J. Beck, P. Schmid, C. Mulatero, N. Miller, S. Dimitrijevic, S. Urva, I. Pylvaenaeinen, K. Petrovic, and B. Johnson, “A phase ib dose-escalation study of everolimus combined with cisplatin and etoposide as first-line therapy in patients with extensive-stage small-cell lung cancer,” Annals of Oncology, vol. 25, pp. 505–511, feb 2014.

40 C. R. Y. Cruz, K. P. Micklethwaite, B. Savoldo, C. A. Ramos, S. Lam, S. Ku, O. Diouf, E. Liu, A. J. Barrett, S. Ito, E. J. Shpall, R. A. Krance, R. T. Kamble, G. Carrum, C. M. Hosing, A. P. Gee, Z. Mei, B. J. Grilley, H. E. Heslop, C. M. Rooney, M. K. Brenner, C. M. Bollard, and G. Dotti, “Infusion of donor-derived CD19-redirected virus-specific t cells for b-cell malignancies relapsed after allogeneic stem cell transplant: a phase 1 study,” Blood, vol. 122, pp. 2965–2973, oct 2013.

41 K. Thornton, A. Chen, M. Trucco, P. Shah, B. Wilky, N. Gul, M. Carrera-Haro, M. F. Ferreira, U. Shafique, J. Powell, C. Meyer, and D. Loeb, “A dose-finding study of temsirolimus and liposomal doxorubicin for patients with recurrent and refractory bone and soft tissue sarcoma,” International Journal of Cancer, vol. 133, pp. 997–1005, mar 2013.

42 S. Sharma, J. Beck, M. Mita, S. Paul, M. M. Woo, M. Squier, B. Gadbaw, and H. M. Prince, “A phase i dose-escalation study of intravenous panobinostat in patients with lymphoma and solid tumors,” Investigational New Drugs, vol. 31, pp. 974–985, feb 2013.

43 C. Sessa, G. I. Shapiro, K. N. Bhalla, C. Britten, K. S. Jacks, M. Mita, V. Papadimitrakopoulou, T. Pluard, T. A. Samuel, M. Akimov, C. Quadt, C. Fernandez-Ibarra, H. Lu, S. Bailey, S. Chica, and U. Banerji, “First-inhuman phase i dose-escalation study of the HSP90 inhibitor AUY922 in patients with advanced solid tumors,” Clinical Cancer Research, vol. 19, pp. 3671–3680, jun 2013.

44 A. F. Schott, M. D. Landis, G. Dontu, K. A. Griffith, R. M. Layman, I. Krop, L. A. Paskett, H. Wong, L. E. Dobrolecki, M. T. Lewis, A. M. Froehlich, J. Paranilam, D. F. Hayes, M. S. Wicha, and J. C. Chang, “Preclinical and clinical studies of gamma secretase inhibitors with docetaxel on human breast tumors,” Clinical Cancer Research, vol. 19, pp. 1512–1524, jan 2013.

45 S. Mangiacavalli, L. Pochintesta, C. Pascutto, F. Cocito, M. Cazzola, and A. Corso, “Good clinical activity and favorable toxicity profile of once weekly bortezomib, fotemustine, and dexamethasone (b-MuD) for the treatment of relapsed multiple myeloma,” American Journal of Hematology, vol. 88, pp. 102–106, dec 2012.

46 A. Larocca, V. Montefusco, S. Bringhen, D. Rossi, C. Crippa, R. Mina, M. Galli, M. Marcatti, G. L. Verde, N. Giuliani, V. Magarotto, T. Guglielmelli, D. Rota-Scalabrini, P. Omedé, A. Santagostino, I. Baldi, A. M. Carella, M. Boccadoro, P. Corradini, and A. Palumbo, “Pomalidomide, cyclophosphamide, and prednisone for relapsed/refractory multiple myeloma: a multicenter phase 1/2 open-label study,” Blood, vol. 122, pp. 2799–2806, oct 2013.

47 R. D. Harvey, T. K. Owonikoko, C. M. Lewis, A. Akintayo, Z. Chen, M. Tighiouart, S. S. Ramalingam, M. P. Fanucchi, P. Nadella, A. Rogatko, D. M. Shin, B. El-Rayes, F. R. Khuri, and J. S. Kauh, “A phase 1 bayesian dose selection study of bortezomib and sunitinib in patients with refractory solid tumor malignancies,” British Journal of Cancer, vol. 108, pp. 762–765, jan 2013.

48 R. S. Finn, R. T. Poon, T. Yau, H.-J. Klümpen, L.-T. Chen, Y.-K. Kang, T.-Y. Kim, C. Gomez-Martin, C. Rodriguez-Lope, T. Kunz, T. Paquet, U. Brandt, D. Sellami, and J. Bruix, “Phase i study investigating everolimus combined with sorafenib in patients with advanced hepatocellular carcinoma,” Journal of Hepatology, vol. 59, pp. 1271–1277, dec 2013.

49 D. J. DeAngelo, A. Spencer, K. N. Bhalla, H. M. Prince, T. Fischer, T. Kindler, F. J. Giles, J. W. Scott, K. Parker, A. Liu, M. Woo, P. Atadja, K. K. Mishra, and O. G. Ottmann, “Phase ia/II, two-arm, open-label, dose-escalation study of oral panobinostat administered via two dosing schedules in patients with advanced hematologic malignancies,” Leukemia, vol. 27, pp. 1628–1636, feb 2013.

50 A. Chiappella, A. Tucci, A. Castellino, V. Pavone, I. Baldi, A. M. Carella, L. Orsucci, M. Zanni, F. Salvi, A. M. Liberati, G. Gaidano, C. Bottelli, B. Rossini, S. Perticone, P. D. Masi, M. Ladetto, G. Ciccone, A. Palumbo, G. Rossi, and U. V. and, “Lenalidomide plus cyclophosphamide, doxorubicin, vincristine, prednisone and rituximab is safe and effective in untreated, elderly patients with diffuse large b-cell lymphoma: a phase i study by the fondazione italiana linfomi,” Haematologica, vol. 98, pp. 1732–1738, jun 2013.

51 D. M. Cannon, M. P. Mehta, J. B. Adkison, D. Khuntia, A. M. Traynor, W. A. Tomé, R. J. Chappell, R. Tolakanahalli, P. Mohindra, S. M. Bentzen, and G. M. Cannon, “Dose-limiting toxicity after hypofractionated dose-escalated radiotherapy in non–small-cell lung cancer,” Journal of Clinical Oncology, vol. 31, pp. 4343–4348, dec 2013.

52 E. Angevin, J. A. Lopez-Martin, C.-C. Lin, J. E. Gschwend, A. Harzstark, D. Castellano, J.-C. Soria, P. Sen, J. Chang, M. Shi, A. Kay, and B. Escudier, “Phase i study of dovitinib (TKI258), an oral FGFR, VEGFR, and PDGFR inhibitor, in advanced or metastatic renal cell carcinoma,” Clinical Cancer Research, vol. 19, pp. 1257–1268, jan 2013.

53 M. C. Foster, C. Amin, P. M. Voorhees, H. W. van Deventer, K. L. Richards, A. Ivanova, J. Whitman, W. M. Chiu, N. D. Barr, and T. Shea, “A phase i dose-escalation study of clofarabine in combination with fractionated gemtuzumab ozogamicin in patients with refractory or relapsed acute myeloid leukemia,” Leukemia & Lymphoma, vol. 53, pp. 1331–1337, jan 2012.

54 C. I. Tsien, D. Brown, D. Normolle, M. Schipper, M. Piert, L. Junck, J. Heth, D. Gomez-Hassan, R. K. T. Haken, T. Chenevert, Y. Cao, and T. Lawrence, “Concurrent temozolomide and dose-escalated intensity-modulated radiation therapy in newly diagnosed glioblastoma,” Clinical Cancer Research, vol. 18, pp. 273–279, nov 2011.

55 A. Tevaarwerk, G. Wilding, J. Eickhoff, R. Chappell, C. Sidor, J. Arnott, H. Bailey, W. Schelman, and G. Liu, “Phase i study of continuous MKC-1 in patients with advanced or metastatic solid malignancies using the modified time-to-event continual reassessment method (TITE-CRM) dose escalation design,” Investigational New Drugs, vol. 30, pp. 1039–1045, jan 2011.

56 R. Sinha, J. L. Kaufman, H. J. Khoury, N. King, P. J. Shenoy, C. Lewis, K. Bumpers, A. Hutchison-Rzepka, M. Tighiouart, L. T. Heffner, M. J. Lechowicz, S. Lonial, and C. R. Flowers, “A phase 1 dose escalation study of bortezomib combined with rituximab, cyclophosphamide, doxorubicin, modified vincristine, and prednisone for untreated follicular lymphoma and other low-grade b-cell lymphomas,” Cancer, vol. 118, pp. 3538–3548, jan 2012.

57 B. J. Schneider, G. P. Kalemkerian, D. Bradley, D. C. Smith, M. J. Egorin, S. Daignault, R. Dunn, and M. Hussain, “Phase i study of vorinostat (suberoylanilide hydroxamic acid, NSC 701852) in combination with docetaxel in patients with advanced and relapsed solid malignancies,” Investigational New Drugs, vol. 30, pp. 249–257, aug 2010.

58 A. W. Roberts, J. F. Seymour, J. R. Brown, W. G. Wierda, T. J. Kipps, S. L. Khaw, D. A. Carney, S. Z. He, D. C. Huang, H. Xiong, Y. Cui, T. A. Busman, E. M. McKeegan, A. P. Krivoshik, S. H. Enschede, and R. Humerickhouse, “Substantial susceptibility of chronic lymphocytic leukemia to BCL2 inhibition: Results of a phase i study of navitoclax in patients with relapsed or refractory disease,” Journal of Clinical Oncology, vol. 30, pp. 488–496, feb 2012.

59 D. A. Reardon, J. J. Vredenburgh, A. Desjardins, K. B. Peters, S. Sathornsumetee, S. Threatt, J. H. Sampson, J. E. Herndon, A. Coan, F. McSherry, J. N. Rich, R. E. McLendon, S. Zhang, and H. S. Friedman, “Phase 1 trial of dasatinib plus erlotinib in adults with recurrent malignant glioma,” Journal of Neuro-Oncology, vol. 108, pp. 499–506, mar 2012.

60 S. Moulder, G. Gladish, J. Ensor, A. M. Gonzalez-Angulo, M. Cristofanilli, J. L. Murray, D. Booser, S. H. Giordano, A. Brewster, J. Moore, E. Rivera, G. N. Hortobagyi, and H. T. Tran, “A phase 1 study of weekly everolimus (RAD001) in combination with docetaxel in patients with metastatic breast cancer,” Cancer, vol. 118, pp. 2378–2384, oct 2011.

61 T. Mazard, M. Ychou, S. Thezenas, S. Poujol, F. Pinguet, A. Thirion, J. P. Bleuse, F. Portales, E. Samalin, and E. Assenat, “Feasibility of biweekly combination chemotherapy with capecitabine, irinotecan, and oxaliplatin in patients with metastatic solid tumors: results of a two-step phase i trial: XELIRI and XELIRINOX,” Cancer Chemotherapy and Pharmacology, vol. 69, pp. 807–814, oct 2011.

62 B. Markman, J. Tabernero, I. Krop, G. Shapiro, L. Siu, L. Chen, M. Mita, M. M. Cuero, S. Stutvoet, D. Birle, O. Anak, W. Hackl, and J. Baselga, “Phase i safety, pharmacokinetic, and pharmacodynamic study of the oral phosphatidylinositol-3-kinase and mTOR inhibitor BGT226 in patients with advanced solid tumors,” Annals of Oncology, vol. 23, pp. 2399–2408, sep 2012.

63 C. Lu, D. J. Stewart, J. J. Lee, L. Ji, R. Ramesh, G. Jayachandran, M. I. Nunez, I. I. Wistuba, J. J. Erasmus, M. E. Hicks, E. A. Grimm, J. M. Reuben, V. Baladandayuthapani, N. S. Templeton, J. D. McMannis, and J. A. Roth, “Phase i clinical trial of systemically administered TUSC2(FUS1)-nanoparticles mediating functional gene transfer in humans,” PLoS ONE, vol. 7, p. e34833, apr 2012.

64 M. Kawahara, A. Kubo, K. Komuta, Y. Fujita, Y. Sasaki, M. Fukushima, T. Daimon, K. Furuse, M. Mishima, and T. Mio, “A phase i study of amrubicin and fixed dose of irinotecan (CPT-11) in relapsed small cell lung cancer: Japan multinational trial organization LC0303,” Journal of Thoracic Oncology, vol. 7, pp. 1845–1849, dec 2012.

65 D. R. Jones, C. A. Moskaluk, H. H. Gillenwater, G. R. Petroni, S. G. Burks, J. Philips, P. K. Rehm, J. Olazagasti, B. D. Kozower, and Y. Bao, “Phase i trial of induction histone deacetylase and proteasome inhibition followed by surgery in non–small-cell lung cancer,” Journal of Thoracic Oncology, vol. 7, pp. 1683–1690, nov 2012.

66 A. J. Jakubowiak, D. Dytfeld, K. A. Griffith, D. Lebovic, D. H. Vesole, S. Jagannath, A. Al-Zoubi, T. Anderson, B. Nordgren, K. Detweiler-Short, K. Stockerl-Goldstein, A. Ahmed, T. Jobkar, D. E. Durecki, K. McDonnell, M. Mietzel, D. Couriel, M. Kaminski, and R. Vij, “A phase 1/2 study of carfilzomib in combination with lenalidomide and low-dose dexamethasone as a frontline treatment for multiple myeloma,” Blood, vol. 120, pp. 1801–1809, aug 2012.

67 K. Geletneky, J. Huesing, J. Rommelaere, J. R. Schlehofer, B. Leuchs, M. Dahm, O. Krebs, M. von Knebel Doeberitz, B. Huber, and J. Hajda, “Phase i/IIa study of intratumoral/intracerebral or intravenous/intracerebral administration of parvovirus h-1 (ParvOryx) in patients with progressive primary or recurrent glioblastoma multiforme: ParvOryx01 protocol,” BMC Cancer, vol. 12, mar 2012.

68 M. Feng, D. E. Smith, D. P. Normolle, J. A. Knol, C. C. Pan, E. Ben-Josef, Z. Lu, M. R. Feng, J. Chen, W. Ensminger, and T. S. Lawrence, “A phase i clinical and pharmacology study using amifostine as a radioprotector in dose-escalated whole liver radiation therapy,” International Journal of Radiation Oncology BiologyPhysics, vol. 83, pp. 1441–1447, aug 2012.

69 M. Farid, B. Chowbay, X. Chen, S. H. Tan, S. Ramasamy, W. H. Koo, H. C. Toh, S. P. Choo, and S. Y. K. Ong, “Phase i pharmacokinetic study of chronomodulated dose-intensified combination of capecitabine and oxaliplatin (XELOX) in metastatic colorectal cancer,” Cancer Chemotherapy and Pharmacology, vol. 70, pp. 141–150, may 2012.

70 K. D. Crew, P. Brown, H. Greenlee, T. B. Bevers, B. Arun, C. Hudis, H. L. McArthur, J. Chang, M. Rimawi, L. Vornik, T. L. Cornelison, A. Wang, H. Hibshoosh, A. Ahmed, M. B. Terry, R. M. Santella, S. M. Lippman, and D. L. Hershman, “Phase IB randomized, double-blinded, placebo-controlled, dose escalation study of polyphenon e in women with hormone receptor-negative breast cancer,” Cancer Prevention Research, vol. 5, pp. 1144–1154, jul 2012.

71 J. C. Bendell, J. Rodon, H. A. Burris, M. de Jonge, J. Verweij, D. Birle, D. Demanse, S. S. D. Buck, Q. C. Ru, M. Peters, M. Goldbrunner, and J. Baselga, “Phase i, dose-escalation study of BKM120, an oral pan-class i PI3k inhibitor, in patients with advanced solid tumors,” Journal of Clinical Oncology, vol. 30, pp. 282–290, jan 2012.

72 E. Ben-Josef, M. Schipper, I. R. Francis, S. Hadley, R. Ten-Haken, T. Lawrence, D. Normolle, D. M. Simeone, C. Sonnenday, R. Abrams, W. Leslie, G. Khan, and M. M. Zalupski, “A phase i/II trial of intensity modulated radiation (IMRT) dose escalation with concurrent fixed-dose rate gemcitabine (FDR-g) in patients with unresectable pancreatic cancer,” International Journal of Radiation Oncology Biology Physics, vol. 84, pp. 1166–1171, dec 2012.

73 A. D. Stasi, S.-K. Tey, G. Dotti, Y. Fujita, A. Kennedy-Nasser, C. Martinez, K. Straathof, E. Liu, A. G. Durett, B. Grilley, H. Liu, C. R. Cruz, B. Savoldo, A. P. Gee, J. Schindler, R. A. Krance, H. E. Heslop, D. M. Spencer, C. M. Rooney, and M. K. Brenner, “Inducible apoptosis as a safety switch for adoptive cell therapy,” New England Journal of Medicine, vol. 365, pp. 1673–1683, nov 2011.

74 K. E. Warren, S. Goldman, I. F. Pollack, J. Fangusaro, P. Schaiquevich, C. F. Stewart, D. Wallace, S. M. Blaney, R. Packer, T. MacDonald, R. Jakacki, J. M. Boyett, and L. E. Kun, “Phase i trial of lenalidomide in pediatric patients with recurrent, refractory, or progressive primary CNS tumors: Pediatric brain tumor consortium study PBTC-018,” Journal of Clinical Oncology, vol. 29, pp. 324–329, jan 2011.

75 J. Vansteenkiste, B. Solomon, M. Boyer, J. Wolf, N. Miller, L. D. Scala, I. Pylvaenaeinen, K. Petrovic, S. Dimitrijevic, B. Anrys, and E. Laack, “Everolimus in combination with pemetrexed in patients with advanced non-small cell lung cancer previously treated with chemotherapy: A phase i study using a novel, adaptive bayesian dose-escalation model,” Journal of Thoracic Oncology, vol. 6, pp. 2120–2129, dec 2011.

76 S. Terakura, Y. Atsuta, M. Sawa, H. Ohashi, T. Kato, S. Nishiwaki, N. Imahashi, T. Yasuda, M. Murata, K. Miyamura, R. Suzuki, T. Naoe, T. Ito, and Y. Morishita, “A prospective dose-finding trial using a modified continual reassessment method for optimization of fludarabine plus melphalan conditioning for marrow transplantation from unrelated donors in patients with hematopoietic malignancies,” Annals of Oncology, vol. 22, pp. 1865–1871, aug 2011.

77 B. D. Smith, R. J. Jones, E. Cho, J. Kowalski, J. E. Karp, S. D. Gore, M. Vala, B. Meade, S. D. Baker, M. Zhao, S. Piantadosi, Z. Zhang, G. Blumenthal, E. D. Warlick, R. A. Brodsky, A. Murgo, M. A. Rudek, and W. H. Matsui, “Differentiation therapy in poor risk myeloid malignancies: Results of a dose finding study of the combination bryostatin-1 and GM-CSF,” Leukemia Research, vol. 35, pp. 87–94, jan 2011.

78 T. Satoh, T. Ura, Y. Yamada, K. Yamazaki, T. Tsujinaka, M. Munakata, T. Nishina, S. Okamura, T. Esaki, Y. Sasaki, W. Koizumi, Y. Kakeji, N. Ishizuka, I. Hyodo, and Y. Sakata, “Genotype-directed, dose-finding study of irinotecan in cancer patients with UGT1a1*28 and/or UGT1a1*6 polymorphisms,” Cancer Science, vol. 102, pp. 1868–1873, aug 2011.

79 J. M. Mehnert, A. R. Tan, R. Moss, E. Poplin, M. N. Stein, M. Sovak, K. Levinson, H. Lin, M. Kane, M. Gounder, Y. Lin, W. J. Shih, E. White, E. H. Rubin, and V. Karantza, “Rationally designed treatment for solid tumors with MAPK pathway activation: A phase i study of paclitaxel and bortezomib using an adaptive dose-finding approach,” Molecular Cancer Therapeutics, vol. 10, pp. 1509–1519, jun 2011.

80 J. Magenau, H. Tobai, A. Pawarode, T. Braun, E. Peres, P. Reddy, C. Kitko, S. Choi, G. Yanik, D. Frame, A. Harris, H. Erba, L. Kujawski, K. Elenitoba-Johnson, J. Sanks, D. Jones, S. Paczesny, J. Ferrara, J. Levine, and S. Mineishi, “Clofarabine and busulfan conditioning facilitates engraftment and provides significant antitumor activity in non-remission hematologic malignancies,” Blood, vol. 118, pp. 4258–4264, oct 2011.

81 S. L. Koolen, P. O. Witteveen, R. S. Jansen, M. H. Langenberg, R. H. Kronemeijer, A. Nol, I. Garcia-Ribas, S. Callies, K. A. Benhadji, C. A. Slapak, J. H. Beijnen, E. E. Voest, and J. H. Schellens, “Phase i study of oral gemcitabine prodrug (LY2334737) alone and in combination with erlotinib in patients with advanced solid tumors,” Clinical Cancer Research, vol. 17, pp. 6071–6082, jul 2011.

82 K. B. Kim, J. Chesney, D. Robinson, H. Gardner, M. M. Shi, and J. M. Kirkwood, “Phase i/II and pharmacodynamic study of dovitinib (TKI258), an inhibitor of fibroblast growth factor receptors and VEGF receptors, in patients with advanced melanoma,” Clinical Cancer Research, vol. 17, pp. 7451–7461, oct 2011.

83 G. Jerusalem, A. Fasolo, V. Dieras, F. Cardoso, J. Bergh, L. Vittori, Y. Zhang, C. Massacesi, T. Sahmoud, and L. Gianni, “Phase i trial of oral mTOR inhibitor everolimus in combination with trastuzumab and vinorelbine in pre-treated patients with HER2-overexpressing metastatic breast cancer,” Breast Cancer Research and Treatment, vol. 125, pp. 447–455, nov 2010.

84 A. J. Jakubowiak, K. A. Griffith, D. E. Reece, C. C. Hofmeister, S. Lonial, T. M. Zimmerman, E. L. Campagnaro, R. L. Schlossman, J. P. Laubach, N. S. Raje, T. Anderson, M. A. Mietzel, C. K. Harvey, S. M. Wear, J. C. Barrickman, C. L. Tendler, D.-L. Esseltine, S. L. Kelley, M. S. Kaminski, K. C. Anderson, and P. G. Richardson, “Lenalidomide, bortezomib, pegylated liposomal doxorubicin, and dexamethasone in newly diagnosed multiple myeloma: a phase 1/2 multiple myeloma research consortium trial,” Blood, vol. 118, pp. 535–543, jul 2011.

85 B. Geoerger, D. Hargrave, F. Thomas, A. Ndiaye, D. Frappaz, F. Andreiuolo, P. Varlet, I. Aerts, R. Riccardi, T. Jaspan, E. Chatelut, M.-C. L. Deley, X. Paoletti, C. Saint-Rose, P. Leblond, B. Morland, J.-C. Gentet, V. Meresse, and G. V. and, “Innovative therapies for children with cancer pediatric phase i study of erlotinib in brainstem glioma and relapsing/refractory brain tumors,” Neuro-Oncology, vol. 13, pp. 109–118, oct 2010.

86 L. Gandhi, D. R. Camidge, M. R. de Oliveira, P. Bonomi, D. Gandara, D. Khaira, C. L. Hann, E. M. McKeegan, E. Litvinovich, P. M. Hemken, C. Dive, S. H. Enschede, C. Nolan, Y.-L. Chiu, T. Busman, H. Xiong, A. P. Krivoshik, R. Humerickhouse, G. I. Shapiro, and C. M. Rudin, “Phase i study of navitoclax (ABT-263), a novel bcl-2 family inhibitor, in patients with small-cell lung cancer and other solid tumors,” Journal of Clinical Oncology, vol. 29, pp. 909–916, mar 2011.

87 M. Fouladi, C. F. Stewart, J. Olson, L. M. Wagner, A. Onar-Thomas, M. Kocak, R. J. Packer, S. Goldman, S. Gururangan, A. Gajjar, T. Demuth, L. E. Kun, J. M. Boyett, and R. J. Gilbertson, “Phase i trial of MK-0752 in children with refractory CNS malignancies: A pediatric brain tumor consortium study,” Journal of Clinical Oncology, vol. 29, pp. 3529–3534, sep 2011.

88 D. Rathkopf, B. Y. Wong, R. W. Ross, A. Anand, E. Tanaka, M. M. Woo, J. Hu, A. Dzik-Jurasz, W. Yang, and H. I. Scher, “A phase i study of oral panobinostat alone and in combination with docetaxel in patients with castration-resistant prostate cancer,” Cancer Chemotherapy and Pharmacology, vol. 66, pp. 181–189, mar 2010.

89 D. M. Peereboom, J. G. Supko, K. A. Carson, T. Batchelor, S. Phuphanich, G. Lesser, T. Mikkelson, J. Fisher, S. Desideri, X. He, and S. A. Grossman, “A phase i/II trial and pharmacokinetic study of ixabepilone in adult patients with recurrent high-grade gliomas,” Journal of Neuro-Oncology, vol. 100, pp. 261–268, may 2010.

90 P. H. O’Donnell, A. S. Artz, S. D. Undevia, R. K. Pai, P. D. Cerro, S. Horowitz, L. A. Godley, J. Hart, F. Innocenti, R. A. Larson, O. M. Odenike, W. Stock, and K. V. Besien, “Phase i study of dose-escalated busulfan with fludarabine and alemtuzumab as conditioning for allogeneic hematopoietic stem cell transplant: reduced clearance at high doses and occurrence of late sinusoidal obstruction syndrome/veno-occlusive disease,” Leukemia & Lymphoma, vol. 51, pp. 2240–2249, oct 2010.

91 R. R. Furman, P. Martin, J. Ruan, Y.-K. K. Cheung, J. M. Vose, A. S. LaCasce, R. Elstrom, M. Coleman, and J. P. Leonard, “Phase 1 trial of bortezomib plus r-CHOP in previously untreated patients with aggressive non-hodgkin lymphoma,” Cancer, vol. 116, pp. 5432–5439, jul 2010.

92 M. Fouladi, C. F. Stewart, S. M. Blaney, A. Onar-Thomas, P. Schaiquevich, R. J. Packer, A. Gajjar, L. E. Kun, J. M. Boyett, and R. J. Gilbertson, “Phase i trial of lapatinib in children with refractory CNS malignancies: A pediatric brain tumor consortium study,” Journal of Clinical Oncology, vol. 28, pp. 4221–4227, sep 2010.

93 F. Andre, M. Campone, R. O’Regan, C. Manlius, C. Massacesi, S. Tarek, P. Mukhopadhyay, J.-C. Soria, M. Naughton, and S. A. Hurvitz, “Phase i study of everolimus plus weekly paclitaxel and trastuzumab in patients with metastatic breast cancer pretreated with trastuzumab,” Journal of Clinical Oncology, vol. 28, pp. 5110–5115, dec 2010.

94 D. M. Loeb, E. Garrett-Mayer, R. F. Hobbs, A. R. Prideaux, G. Sgouros, O. Shokek, M. D. Wharam, T. Scott, and C. L. Schwartz, “Dose-finding study of153sm-EDTMP in patients with poor-prognosis osteosarcoma,” Cancer, vol. 115, pp. 2514–2522, jun 2009.

95 B. sheng Li, T. Zhou, Z. tang Wang, H. sheng Li, H. fu Sun, Z. cheng Zhang, H. qun Lin, Y. mei Wei, H. yi Gong, W. Huang, Y. Yi, and L. ying Wang, “Phase i study of concurrent selective lymph node late course accelerated hyper-fractionated radiotherapy and capecitabine, cisplatin for locally advanced esophageal squamous cell carcinoma,” Radiotherapy and Oncology, vol. 93, pp. 458–461, dec 2009.

96 G. D. Demetri, P. L. Russo, I. R. MacPherson, D. Wang, J. A. Morgan, V. G. Brunton, P. Paliwal, S. Agrawal, M. Voi, and T. J. Evans, “Phase i dose-escalation and pharmacokinetic study of dasatinib in patients with advanced solid tumors,” Clinical Cancer Research, vol. 15, pp. 6232–6240, sep 2009.

97 H. Borghaei, K. Alpaugh, G. Hedlund, G. Forsberg, C. Langer, A. Rogatko, R. Hawkins, S. Dueland, U. Lassen, and R. B. Cohen, “Phase i dose escalation, pharmacokinetic and pharmacodynamic study of naptumomab estafenatox alone in patients with advanced cancer and with docetaxel in patients with advanced non–small-cell lung cancer,” Journal of Clinical Oncology, vol. 27, pp. 4116–4123, sep 2009.

98 D. B. Bailey, K. M. Rassnick, N. L. Dykes, and L. Pendyala, “Phase i evaluation of carboplatin by use of a dosing strategy based on a targeted area under the platinum concentration-versus-time curve and individual glomerular filtration rate in cats with tumors,” American Journal of Veterinary Research, vol. 70, pp. 770–776, jun 2009.

99 L. Trümper, C. Zwick, M. Ziepert, K. Hohloch, R. Schmits, M. Mohren, R. Liersch, M. Bentz, U. Graeven, U. Wruck, M. Hoffmann, B. Metzner, D. Hasenclever, M. Loeffler, and M. Pfreundschuh, “Dose-escalated CHOEP for the treatment of young patients with aggressive non-hodgkin’s lymphoma: I. a randomized dose escalation and feasibility study with bi-and tri-weekly regimens,” Annals of Oncology, vol. 19, pp. 538–544, mar 2008.

100 S. Saji, M. Toi, S. Morita, H. Iwata, Y. Ito, S. Ohno, T. Kobayashi, Y. Hozumi, and J. Sakamoto, “Dose-finding phase i and pharmacokinetic study of capecitabine (xeloda) in combination with epirubicin and cyclophosphamide (CEX) in patients with inoperable or metastatic breast cancer,” Oncology, vol. 72, no. 5–6, pp. 330–337, 2007.

101 S. Rao, N. Starling, D. Cunningham, M. Benson, A. Wotherspoon, C. Lüpfert, R. Kurek, J. Oates, J. Baselga, and A. Hill, “Phase i study of epirubicin, cisplatin and capecitabine plus matuzumab in previously untreated patients with advanced oesophagogastric cancer,” British Journal of Cancer, vol. 99, pp. 868–874, sep 2008.

102 B. Neuenschwander, M. Branson, and T. Gsponer, “Critical aspects of the bayesian approach to phase i cancer trials,” Statistics in Medicine, vol. 27, no. 13, pp. 2420–2439, 2008.

103 T. J. MacDonald, C. F. Stewart, M. Kocak, S. Goldman, R. G. Ellenbogen, P. Phillips, D. Lafond, T. Y. Poussaint, M. W. Kieran, J. M. Boyett, and L. E. Kun, “Phase i clinical trial of cilengitide in children with refractory brain tumors: Pediatric brain tumor consortium study PBTC-012,” Journal of Clinical Oncology, vol. 26, pp. 919–924, feb 2008.

104 A. Jimeno, M. A. Rudek, P. Kulesza, W. W. Ma, J. Wheelhouse, A. Howard, Y. Khan, M. Zhao, H. Jacene, W. A. Messersmith, D. Laheru, R. C. Donehower, E. Garrett-Mayer, S. D. Baker, and M. Hidalgo, “Pharmacodynamic-guided modified continuous reassessment method–based, dose-finding study of rapamycin in adult patients with solid tumors,” Journal of Clinical Oncology, vol. 26, pp. 4172–4179, sep 2008.

105 S. Gururangan, C. D. Turner, C. F. Stewart, M. O’Shaughnessy, M. Kocak, T. Y. Poussaint, P. C. Phillips, S. Goldman, R. Packer, I. F. Pollack, S. M. Blaney, V. Karsten, S. L. Gerson, J. M. Boyett, H. S. Friedman, and L. E. Kun, “Phase i trial of VNP40101m (cloretazine) in children with recurrent brain tumors: A pediatric brain tumor consortium study,” Clinical Cancer Research, vol. 14, pp. 1124–1130, feb 2008.

106 B. Guillot, A. Khamari, D. Cupissol, M. Delaunay, C. Bedane, B. Dreno, M. C. Picot, and O. Dereure, “Temozolomide associated with PEG-interferon in patients with metastatic melanoma: a multicenter prospective phase i/II study,” Melanoma Research, vol. 18, pp. 141–146, apr 2008.

107 S. A. Grossman, K. A. Carson, S. Phuphanich, T. Batchelor, D. Peereboom, L. B. Nabors, G. Lesser, F. Hausheer, and J. G. Supko, “Phase i and pharmacokinetic study of karenitecin in patients with recurrent malignant gliomas,” Neuro-Oncology, vol. 10, pp. 608–616, aug 2008.

108 J. S. de Bono, R. Kristeleit, A. Tolcher, P. Fong, S. Pacey, V. Karavasilis, M. Mita, H. Shaw, P. Workman, S. Kaye, E. K. Rowinsky, W. Aherne, P. Atadja, J. W. Scott, and A. Patnaik, “Phase i pharmacokinetic and pharmacodynamic study of LAQ824, a hydroxamate histone deacetylase inhibitor with a heat shock protein-90 inhibitory profile, in patients with advanced solid tumors,” Clinical Cancer Research, vol. 14, pp. 6663–6673, oct 2008.

109 J. B. Adkison, D. Khuntia, S. M. Bentzen, G. M. Cannon, W. A. Tome, H. Jaradat, W. Walker, A. M. Traynor, T. Weigel, and M. P. Mehta, “Dose escalated, hypofractionated radiotherapy using helical tomotherapy for inoperable non-small cell lung cancer: Preliminary results of a risk-stratified phase i dose escalation study,” Technology in Cancer Research & Treatment, vol. 7, pp. 441–447, dec 2008.

110 F. Atzori, J. Tabernero, A. Cervantes, L. Prudkin, J. Andreu, E. Rodriguez-Braun, A. Domingo, J. Guijarro, C. Gamez, J. Rodon, S. D. Cosimo, H. Brown, J. Clark, J. S. Hardwick, R. A. Beckman, W. D. Hanley, K. Hsu, E. Calvo, S. Rosello, R. B. Langdon, and J. Baselga, “A phase i pharmacokinetic and pharmacodynamic study of dalotuzumab (MK-0646), an anti-insulin-like growth factor-1 receptor monoclonal antibody, in patients with advanced solid tumors,” Clinical Cancer Research, vol. 17, pp. 6304–6312, aug 2011.

111 J. R. Berenson, O. Yellin, R. Patel, H. Duvivier, Y. Nassir, R. Mapes, C. D. Abaya, and R. A. Swift, “A phase i study of samarium lexidronam/bortezomib combination therapy for the treatment of relapsed or refractory multiple myeloma,” Clinical Cancer Research, vol. 15, pp. 1069–1075, feb 2009.

112 T. Conroy, F. Viret, E. François, J. F. Seitz, V. Boige, M. Ducreux, M. Ychou, J. P. Metges, M. Giovannini, Y. Yataghene, and D. Peiffert, “Phase i trial of oxaliplatin with fluorouracil, folinic acid and concurrent radiotherapy for oesophageal cancer,” British Journal of Cancer, vol. 99, pp. 1395–1401, oct 2008.

113 F. L. Day, T. Leong, S. Ngan, R. Thomas, M. Jefford, J. R. Zalcberg, D. Rischin, J. McKendick, A. D. Milner, J. D. Iulio, A. Matera, and M. Michael, “Phase i trial of docetaxel, cisplatin and concurrent radical radiotherapy in locally advanced oesophageal cancer,” British Journal of Cancer, vol. 104, pp. 265–271, dec 2010.

114 M. Dong, Z.-Q. Ning, P.-Y. Xing, J.-L. Xu, H.-X. Cao, G.-F. Dou, Z.-Y. Meng, Y.-K. Shi, X.-P. Lu, and F.-Y. Feng, “Phase i study of chidamide (CS055/HBI-8000), a new histone deacetylase inhibitor, in patients with advanced solid tumors and lymphomas,” Cancer Chemotherapy and Pharmacology, vol. 69, pp. 1413–1422, feb 2012.

115 P. Forsyth, G. Roldán, D. George, C. Wallace, C. A. Palmer, D. Morris, G. Cairncross, M. V. Matthews, J. Markert, Y. Gillespie, M. Coffey, B. Thompson, and M. Hamilton, “A phase i trial of intratumoral administration of reovirus in patients with histologically confirmed recurrent malignant gliomas,” Molecular Therapy, vol. 16, pp. 627–632, mar 2008.

116 A. Frost, K. Mross, S. Steinbild, S. Hedbom, C. Unger, R. Kaiser, D. Trommeshauser, and G. Munzert, “Phase i study of the plk1 inhibitor BI 2536 administered intravenously on three consecutive days in advanced solid tumours,” Current Oncology, vol. 19, feb 2012.

117 I. Gojo, M. Tan, H.-B. Fang, M. Sadowska, R. Lapidus, M. R. Baer, F. Carrier, J. H. Beumer, B. N. Anyang, R. K. Srivastava, et al., “Translational phase i trial of vorinostat (suberoylanilide hydroxamic acid) combined with cytarabine and etoposide in patients with relapsed, refractory, or high-risk acute myeloid leukemia,” Clinical Cancer Research, vol. 19, no. 7, pp. 1838–1851, 2013.

118 H. M. Kantarjian, S. Padmanabhan, W. Stock, M. S. Tallman, G. A. Curt, J. Li, A. Osmukhina, K. Wu, D. Huszar, G. Borthukar, S. Faderl, G. Garcia-Manero, T. Kadia, K. Sankhala, O. Odenike, J. K. Altman, and M. Minden, “Phase i/II multicenter study to assess the safety, tolerability, pharmacokinetics and pharmacodynamics of AZD4877 in patients with refractory acute myeloid leukemia,” Investigational New Drugs, vol. 30, pp. 1107–1115, apr 2011.

119 K. B. Kim, W.-J. Hwu, N. E. Papadopoulos, A. Y. Bedikian, L. H. Camacho, C. Ng, I. M. Hernandez, A. M. Frost, M. A. Jack, and P. Hwu, “Phase i study of the combination of docetaxel, temozolomide and cisplatin in patients with metastatic melanoma,” Cancer Chemotherapy and Pharmacology, vol. 64, pp. 161–167, nov 2008.

120 K. J. Kimball, M. A. Preuss, M. N. Barnes, M. Wang, G. P. Siegal, W. Wan, H. Kuo, S. Saddekni, C. R. Stockard, W. E. Grizzle, R. D. Harris, R. Aurigemma, D. T. Curiel, and R. D. Alvarez, “A phase i study of a tropism-modified conditionally replicative adenovirus for recurrent malignant gynecologic diseases,” Clinical Cancer Research, vol. 16, pp. 5277–5287, oct 2010.

121 P. L. Kunz, A. R. He, A. D. Colevas, M. J. Pishvaian, J. J. Hwang, P. L. Clemens, M. Messina, R. Kaleta, F. Abrahao, B. I. Sikic, and J. L. Marshall, “Phase i trial of ixabepilone administered as three oral doses each separated by 6 hours every 3 weeks in patients with advanced solid tumors,” Investigational New Drugs, vol. 30, pp. 2364–2370, feb 2012.

122 K.-W. Lee, S. R. Park, D.-Y. Oh, Y.-I. Park, R. Khosravan, X. Lin, S.-Y. Lee, E.-J. Roh, O. Valota, M. J. Lechuga, and Y.-J. Bang, “Phase i study of sunitinib plus capecitabine/cisplatin or capecitabine/oxaliplatin in advanced gastric cancer,” Investigational New Drugs, vol. 31, pp. 1547–1558, oct 2013.

123 P. LoRusso, K. Venkatakrishnan, E. G. Chiorean, D. Noe, J.-T. Wu, S. Sankoh, M. Corvez, and E. A. Sausville, “Phase 1 dose-escalation, pharmacokinetic, and cerebrospinal fluid distribution study of TAK-285, an investigational inhibitor of EGFR and HER2,” Investigational New Drugs, vol. 32, pp. 160–170, jul 2013.

124 J. J. Luke, D. R. D’Adamo, M. A. Dickson, M. L. Keohan, R. D. Carvajal, R. G. Maki, E. de Stanchina, E. Musi, S. Singer, and G. K. Schwartz, “The cyclin-dependent kinase inhibitor flavopiridol potentiates doxorubicin efficacy in advanced sarcomas: Preclinical investigations and results of a phase i dose-escalation clinical trial,” Clinical Cancer Research, vol. 18, pp. 2638–2647, feb 2012.

125 M. D. Michaelson, A. X. Zhu, D. P. Ryan, D. F. McDermott, G. I. Shapiro, L. Tye, I. Chen, P. Stephenson, S. Patyna, A. Ruiz-Garcia, and A. B. Schwarzberg, “Sunitinib in combination with gemcitabine for advanced solid tumours: a phase i dose-finding study,” British Journal of Cancer, vol. 108, pp. 1393–1401, mar 2013.

126 M. Mita, K. R. Kelly, A. Mita, A. D. Ricart, O. Romero, A. Tolcher, L. Hook, C. Okereke, I. Krivelevich, D. P. Rossignol, F. J. Giles, E. K. Rowinsky, and C. Takimoto, “Phase i study of e7820, an oral inhibitor of integrin –2 expression with antiangiogenic properties, in patients with advanced malignancies,” Clinical Cancer Research, vol. 17, pp. 193–200, jan 2011.

127 Y. Oki, D. Buglio, M. Fanale, L. Fayad, A. Copeland, J. Romaguera, L. W. Kwak, B. Pro, S. de Castro Faria, S. Neelapu, N. Fowler, F. Hagemeister, J. Zhang, S. Zhou, L. Feng, and A. Younes, “Phase i study of panobinostat plus everolimus in patients with relapsed or refractory lymphoma,” Clinical Cancer Research, vol. 19, pp. 6882–6890, oct 2013.

128 D. A. Pollyea, H. E. Kohrt, L. Gallegos, M. E. Figueroa, O. Abdel-Wahab, B. Zhang, S. Bhattacharya, J. Zehnder, M. Liedtke, J. R. Gotlib, S. Coutre, C. Berube, A. Melnick, R. Levine, B. S. Mitchell, and B. C. Medeiros, “Safety, efficacy and biological predictors of response to sequential azacitidine and lenalidomide for elderly patients with acute myeloid leukemia,” Leukemia, vol. 26, pp. 893–901, oct 2011.

129 A. V. Rao, D. A. Rizzieri, C. M. DeCastro, L. F. Diehl, A. S. Lagoo, J. O. Moore, and J. P. Gockerman, “Phase i study of dose dense induction and consolidation with gemtuzumab ozogamicin and high dose cytarabine in older adults with AML,” Journal of Geriatric Oncology, vol. 3, pp. 220–227, jul 2012.

130 S. L. Sanborn, M. M. Cooney, A. Dowlati, J. M. Brell, S. Krishnamurthi, J. Gibbons, J. A. Bokar, C. Nock, A. Ness, and S. C. Remick, “Phase i trial of docetaxel and thalidomide: a regimen based on metronomic therapeutic principles,” Investigational New Drugs, vol. 26, pp. 355–362, may 2008.

131 G. I. Shapiro, S. McCallum, L. M. Adams, L. Sherman, S. Weller, S. Swann, H. Keer, D. Miles, T. Müller, and P. LoRusso, “A phase 1 dose-escalation study of the safety and pharmacokinetics of once-daily oral foretinib, a multi-kinase inhibitor, in patients with solid tumors,” Investigational New Drugs, vol. 31, pp. 742–750, oct 2012.

132 S. Srivastava, D. Jones, L. L. Wood, J. E. Schwartz, R. P. Nelson, R. Abonour, A. Secrest, E. Cox, J. Baute, C. Sullivan, K. Kane, M. J. Robertson, and S. S. Farag, “A phase i trial of high-dose clofarabine, etoposide, and cyclophosphamide and autologous peripheral blood stem cell transplantation in patients with primary refractory and relapsed and refractory non-hodgkin lymphoma,” Biology of Blood and Marrow Transplantation, vol. 17, pp. 987–994, jul 2011.

133 A. M. Tsimberidou, L. H. Camacho, S. Verstovsek, C. Ng, D. S. Hong, C. K. Uehara, C. Gutierrez, S. Daring, J. Stevens, P. B. Komarnitsky, B. Schwartz, and R. Kurzrock, “A phase i clinical trial of darinaparsin in patients with refractory solid tumors,” Clinical Cancer Research, vol. 15, pp. 4769–4776, jul 2009.

134 K. Yamamoto, K. Kokawa, N. Umesaki, R. Nishimura, K. Hasegawa, I. Konishi, F. Saji, M. Nishida, H. Noguchi, and K. Takizawa, “Phase i study of combination chemotherapy with irinotecan hydrochloride and nedaplatin for cervical squamous cell carcinoma: Japanese gynecologic oncology group study,” Oncology reports, vol. 21, no. 4, pp. 1005–1009, 2009.

135 A. Younes, N. L. Bartlett, J. P. Leonard, D. A. Kennedy, C. M. Lynch, E. L. Sievers, and A. Forero-Torres, “Brentuximab vedotin (sgn-35) for relapsed cd30-positive lymphomas,” New England Journal of Medicine, vol. 363, no. 19, pp. 1812–1821, 2010.

136 J. A. Garcia, T. Mekhail, P. Elson, L. Wood, R. M. Bukowski, R. Dreicer, and B. I. Rini, “Phase i/II trial of subcutaneous interleukin-2, granulocyte-macrophage colony-stimulating factor and interferon-alpha in patients with metastatic renal cell carcinoma,” BJU International, vol. 109, pp. 63–69, jan 2011.

137 B. Saraiya, M. Gounder, J. Dutta, A. Saleem, C. Collazo, L. Zimmerman, A. Nazar, M. Gharibo, D. Schaar, Y. Lin, W. Shih, J. Aisner, R. K. Strair, and E. H. Rubin, “Sequential topoisomerase targeting and analysis of mechanisms of resistance to topotecan in patients with acute myelogenous leukemia,” Anti-Cancer Drugs, vol. 19, pp. 411–420, apr 2008.

138 S. L. Spunt, S. A. Grupp, T. A. Vik, V. M. Santana, D. J. Greenblatt, J. Clancy, A. Berkenblit, M. Krygowski, R. Ananthakrishnan, J. P. Boni, and R. J. Gilbertson, “Phase i study of temsirolimus in pediatric patients with recurrent/refractory solid tumors,” Journal of Clinical Oncology, vol. 29, pp. 2933–2940, jul 2011.

139 K. H. Yu, S. C. H. Yu, E. P. Hui, M. K. M. Kam, A. C. Vlantis, E. Yuen, and A. T. C. Chan, “Accelerated fractionation radiotherapy and late intensification with 2 intra-arterial cisplatin infusions for locally advanced head and neck squamous cell carcinoma,” Head & Neck, pp. NA–NA, 2009.

140 E. A. Eisenhauer, P. Therasse, J. Bogaerts, L. H. Schwartz, D. Sargent, R. Ford, J. Dancey, S. Arbuck, S. Gwyther, M. Mooney, L. Rubinstein, L. Shankar, L. Dodd, R. Kaplan, D. Lacombe, and J. Verweij, “New response evaluation criteria in solid tumours: Revised RECIST guideline (version 1.1),” European Journal of Cancer, vol. 45, no. 2, pp. 228–247, 2009. Publisher: Elsevier Ltd ISBN: 1879–0852 (Electronic).

141 B. D. Cheson, J. M. Bennett, K. J. Kopecky, T. Büchner, C. L. Willman, E. H. Estey, C. A. Schiffer, H. Doehner, M. S. Tallman, T. A. Lister, F. Lo-Coco, R. Willemze, A. Biondi, W. Hiddemann, R. A. Larson, B. Löwenberg, M. A. Sanz, D. R. Head, R. Ohno, C. D. Bloomfield, F. Lo-Cocco, and International Working Group for Diagnosis, Standardization of Response Criteria, Treatment Outcomes, and Reporting Standards for Therapeutic Trials in Acute Myeloid Leukemia, “Revised recommendations of the International Working Group for Diagnosis, Standardization of Response Criteria, Treatment Outcomes, and Reporting Standards for Therapeutic Trials in Acute Myeloid Leukemia,” Journal of Clinical Oncology: Official Journal of the American Society of Clinical Oncology, vol. 21, pp. 4642–4649, Dec. 2003.

142 M. Hallek, B. D. Cheson, D. Catovsky, F. Caligaris-Cappio, G. Dighiero, H. Döhner, P. Hillmen, M. Keating, E. Montserrat, N. Chiorazzi, S. Stilgenbauer, K. R. Rai, J. C. Byrd, B. Eichhorst, S. O’Brien, T. Robak, J. F. Seymour, and T. J. Kipps, “iwCLL guidelines for diagnosis, indications for treatment, response assessment, and supportive management of CLL,” Blood, vol. 131, no. 25, pp. 2745–2760, 2018.

143 K. Brock, V. Homer, G. Soul, C. Potter, C. Chiuzan, and S. Lee, “Dose-level toxicity and efficacy outcomes from dose-finding clinical trials in oncology.” https://doi.org/10.25500/edata.bham.00000337, 2019.

144 J. Macdougall, “Analysis of Dose–Response Studies—Emax Model,” in Dose Finding in Drug Development, Springer New York, 2006.

145 B. Bornkamp, DoseFinding: Planning and Analyzing Dose Finding Experiments, 2019. R package version 0.9–17.

146 P.-C. Bürkner, “Advanced Bayesian multilevel modeling with the R package brms,” The R Journal, vol. 10, no. 1, pp. 395–411, 2018.

147 R Core Team, R: A Language and Environment for Statistical Computing. R Foundation for Statistical Computing, Vienna, Austria, 2020.

148 H. Wickham, M. Averick, J. Bryan, W. Chang, L. D. McGowan, R. François, G. Grolemund, A. Hayes, L. Henry, J. Hester, M. Kuhn, T. L. Pedersen, E. Miller, S. M. Bache, K. Müller, J. Ooms, D. Robinson, D. P. Seidel, V. Spinu, K. Takahashi, D. Vaughan, C. Wilke, K. Woo, and H. Yutani, “Welcome to the tidyverse,” Journal of Open Source Software, vol. 4, no. 43, p. 1686, 2019.

149 M. Kay, tidybayes: Tidy Data and Geoms for Bayesian Models, 2020. R package version 2.0.3.

150 H. Wickham, ggplot2: Elegant Graphics for Data Analysis. Springer-Verlag New York, 2016.

151 S. M. Lee, D. Backenroth, Y. K. K. Cheung, D. L. Hershman, D. Vulih, B. Anderson, P. Ivy, and L. Minasian, “Case example of dose optimization using data from bortezomib dose-finding clinical trials,” Journal of Clinical Oncology, vol. 34, no. 12, p. 1395, 2016.

152 S. Zohar, I. Baldi, G. Forni, F. Merletti, G. Masucci, and D. Gregori, “Planning a bayesian early-phase phase i/ii study for human vaccines in her2 carcinomas,” Pharmaceutical statistics, vol. 10, no. 3, pp. 218–226, 2011.

153 A. Hazim, G. Mills, V. Prasad, A. Haslam, and E. Y. Chen, “Relationship Between Response and Dose in Published, Contemporary Phase I Oncology Trials,” Journal of the National Comprehensive Cancer Network: JNCCN, vol. 18, no. 4, pp. 428–433, 2020.

